# The role of climate and immunity in Brazil’s unprecedented outbreak of dengue in 2024

**DOI:** 10.64898/2026.10.31.26366870

**Authors:** Bachir Assao Neino, Bernardo García-Carreras, Mondal Hasan Zahid, Angkana T. Huang, Youngsuk Ko, Paloma Cárcamo, Luciano A. Moreira, Mauricio L. Nogueira, Mauro M. Teixeira, Albert I. Ko, Matt D.T. Hitchings, Derek A.T. Cummings

## Abstract

Brazil experienced a large dengue epidemic during the 2024 season, with almost 5.7 million total cases (compared to under 1.3 million in 2023), raising questions about underlying drivers, including rising temperatures, expansion into susceptible populations, changes in reporting, and novel strains. We compared state-level 2024 incidence with 2000–2023 data to quantify anomalies. Nine of 27 states, primarily in the Southeast, recorded their highest incidence in 25 years, accounting for 97% of cases while comprising only 67% of the population. In contrast, nine states (mostly in the North and Northeast) reported below-average incidence. Counterintuitively, the correlation between state-level temperature and incidence declined significantly over the study period, turning negative by 2024 (*r* = −0.72), a reversal from the positive correlations observed at the start. To determine whether this pattern was driven by northern temperatures being too high for transmission, or by spatial variation in immunity, we fit generalized additive models to effective reproductive numbers. Transmission remained substantial at the highest temperatures, while reproductive numbers declined with lagged cumulative incidences, indicating susceptibility depletion was important. We then estimated state-level susceptibility using a mechanistic SEIR-SEI model, describing a trade-off: high temperatures supported high transmission rates, but were associated with reduced susceptibility. Linear regression models confirmed both temperature and susceptibility were necessary to predict the 2024 outbreaks, suggesting that immunity plays an important role in modulating temperature’s effect on incidence. Future surveillance integrating measures of immunity could enhance forecasts and early-warning systems, particularly as climate change expands dengue into immunologically naïve populations.

**Significance Statement:** Brazil experienced an unprecedented dengue epidemic in 2024, with nearly 5.7 million reported cases. The relative role of high temperature and population immunity in driving this large outbreak remains poorly understood. High temperatures are known to support increased transmission but surprisingly, the relationship between incidence and temperature had turned negative by this outbreak. Our results show that transmission remained viable at the highest temperatures, but outbreaks were constrained by immunity acquired through prior infection. Large epidemics occurred only where favorable temperatures coincided with high remaining susceptibility, itself largely a legacy of past, temperature-driven transmission. These results demonstrate that immunity modulates climate-driven dengue risk, highlighting the value of estimating population immunity alongside climate.

## Introduction

The number of reported dengue cases worldwide more than doubled from 2023 to 2024^1^. This increase was in part driven by the largest epidemic ever recorded in the Americas with Brazil reporting almost 5.7 million suspected cases, representing 79% of the total in the Americas. Transmission is endemic in large parts of Brazil and large-scale outbreaks have occurred regularly; however, the 2024 outbreak was unprecedented.

Of all hypotheses being proposed to explain Brazil’s unprecedented 2024 dengue outbreak, climate change is the most common. An El Niño event increased temperatures and altered rainfall, increasing mosquito density and expanding the vector’s range^1,2^. Temperature shapes dengue transmission through its effect on vector life-history traits and viral replication^3^. Mechanistic models based on mosquito thermal physiology predict that basic reproductive numbers (*R*_0_) increase nonmonotonically with temperature, reaching a peak before declining at higher temperatures^3,4^. These temperature-dependent *R*_0_ curves have been used to project future expansion of suitable habitat and epidemic potential under climate change^3,5,6^. More recently, a global analysis found dengue incidence to similarly depend nonlinearly on temperature^7^. The nonlinear effects of temperature, combined with the multiplicative nature of transmission dynamics, means that even modest increases in temperature could lead to disproportionate case surges.

In addition to climate, other contributing factors, many related to immunity, have been put forward. Serotype dynamics may have created local gaps in immunity: the co-circulation of multiple serotypes^8^, the re-emergence of DENV-3 in São Paulo^8–10^, and the DENV-2 Cosmopolitan genotype into the Western Amazon^11^ all placed populations with limited serotype-specific immunity at greater risk. Prior heterotypic immunity may have enhanced disease severity through antibody-dependent enhancement, potentially inflating case counts^12^. Increased human mobility may also have facilitated introduction into new areas. Urbanization and deforestation in the Central-West and Southeast may have expanded *Aedes aegypti* breeding sites^13,14^, exposing populations with low immunity. Other factors unrelated to immunity have also been proposed: co-circulating arboviruses with similar symptoms (e.g., Oropouche, chikungunya, and Zika) have posed challenges to surveillance and likely inflated suspected dengue counts^15–18^, while differences in reporting may have contributed to the elevated counts observed in 2024.

Climate change and immunity are often examined in isolation, with less attention given to how they may interact spatially. As climates warm, areas transition from unsuitable to suitable for sustained transmission, exposing fully susceptible populations to dengue. As seen in pandemics, initial outbreaks of immunizing pathogens in such populations greatly exceed subsequent outbreaks, as initial large outbreaks induce immunity that reduces or blunts future transmission. A recent exploratory analysis of municipality-level dengue incidence and temperature in Brazil observed a shift from a positive correlation in 2013 to a negative correlation in 2024, with population immunity proposed as a potential driver of this change.^19^ Other authors have highlighted the expansion of dengue into areas with low historical transmission as possible drivers of incidence in recent years^20,21^, but often the focus is on the impact of temperature on mosquito abundance and transmission alone^1,2,22–24^. Quantifying susceptibility from case data alone remains challenging: widespread asymptomatic infection^25^ and multiple circulating serotypes and genotypes^26^, and limited serological surveillance obscure real immunity levels, making it difficult to disentangle temperature’s direct effect on transmission from immunity-driven blunting of transmission driven by historical transmission that itself is temperature driven.

To investigate the drivers of Brazil’s 2024 dengue epidemic, we analyzed 24 years of state and municipality-level case and temperature data. A striking pattern observed in Brazil requires explanation: many northern states with consistently high temperatures (previously associated with higher dengue transmission) experienced average or below-average case counts, while southeastern states with comparatively lower temperatures, saw unprecedented outbreaks. We fit models to quantify the importance of temperature and population immunity in explaining the 2024 outbreak and its spatial patterns. Using a mechanistic, temperature-dependent transmission model, we inferred state-level susceptibility indices, and tested whether accounting for spatial variation in immunity could resolve the apparent contradiction between temperature and incidence patterns. Our study addresses how climate-driven range expansion interacts with immunity dynamics to shape dengue emergence and outbreaks, insights that have implications for anticipating future outbreaks as warming continues.

## Results

### Temperature is insufficient to explain the spatial patterns in the 2024 outbreak

The Brazil 2024 dengue season (defined as July 2023 to June 2024) was unprecedented in scale (Fig.1A), with weekly case counts exceeding all prior seasons in our dataset. Spatially, the burden was concentrated in the Southeast and South (Fig. 1C); nine states, primarily in the Southeast, accounted for 97% of cases despite comprising 67% of the population. This concentration is a reversal of historic spatial patterns: the heatmap of case distribution relative to population (Fig. 1B) shows that states in the Southeast and South - historically less dominant - were disproportionately represented in 2024, while many traditional hotspots in the Northeast and North contributed relatively less compared with their historical profiles. Equivalent descriptive results using confirmed cases only are shown in SI Appendix, Fig. S1.

**Figure 1.**
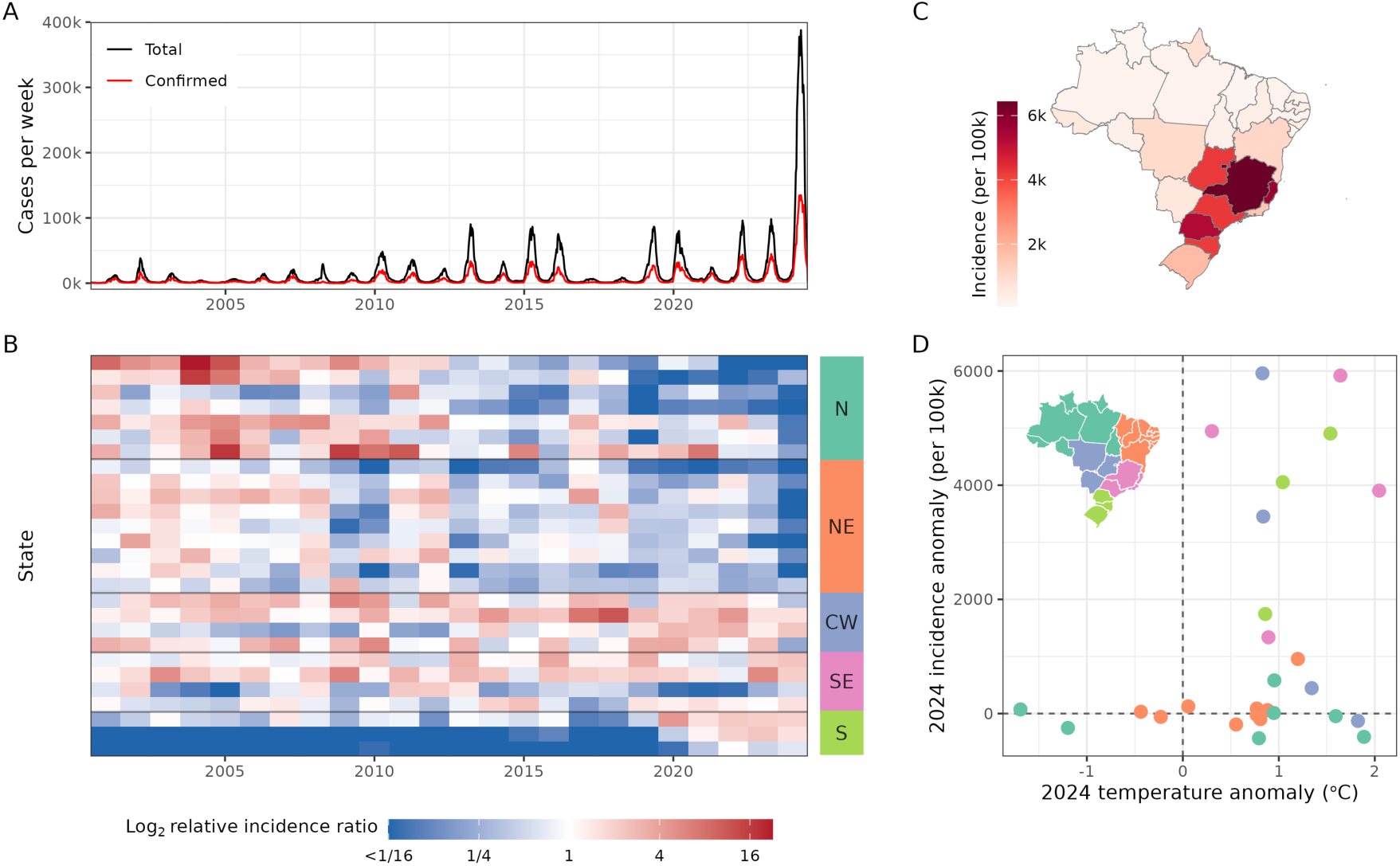
Spatiotemporal patterns and drivers of the 2024 dengue outbreak in Brazil. (A) Weekly dengue cases in Brazil from 2001-2024, showing the unprecedented 2024 outbreak. (B) Log_2_ relative incidence ratios by state and season, calculated as the proportion of national total cases reported by each state, divided by the proportion of the national population in each state. Red indicates states contributing more cases than expected given their population share, revealing a southern shift in 2024. States are ordered geographically by region and latitude, from north (top) to south (bottom); the regions are indicated to the right. Values on the low end of the scale were capped at log_2_ 1/16 for clarity. (C) Annual incidence rates (per 100,000, based on total cases) by state for the 2024 season, showing the disproportionate burden in southern and southeastern regions. (D) Relationship between temperature anomalies and incidence (per 100,000) anomalies by state for the 2024 season. Dashed lines, for anomalies equal to zero, represent the means for the 2001 to 2023 seasons. The points are colored by region, shown in the inset map. See SI Appendix, Fig. S1 for the equivalent figure using confirmed cases only.

The geographic shift in incidence toward cooler southern states raises the question of whether those same states experienced unusually higher temperatures relative to the rest of the country. Most states recorded higher-than-average temperatures in 2024, with only four exceptions (two in the North and two in the Northeast; Fig. 1D). We next systematically explored the relationship between temperature and incidence by calculating the Pearson correlation (adjusted for spatial autocorrelation) between log_10_ total incidence and mean temperature for each of the seasons 2001-2024. The Pearson correlation for the 2024 season was −0.72 (95% confidence intervals (CI): −0.96 to 0.09; Fig. 2), a large negative point estimate indicating that states with a lower mean temperature tended to have larger outbreaks in 2024, though the association is not statistically significant. Correlations prior to the mid-2010s tended to be positive, and were significantly positive for the 2001, 2003-2009, 2011, 2012, and 2017 seasons (as per Pearson correlation coefficients; Fig. 2). We found a significant declining trend in the correlation over the study period, fitted on the Fisher-*z* scale (Methods): −0.059 per year for Pearson (95% CI: −0.082 to −0.035) and −0.044 per year for Spearman (95% CI: −0.062 to −0.026); the fitted trend crosses zero in the mid-2010s and its 95% band excludes zero over the most recent seasons. The corresponding confirmed-case correlations are shown in SI Appendix, Fig. S2. The same declining pattern was observed at different spatial resolutions (health regions and municipalities; see SI Appendix S4 and Fig. S3). This decline suggests that temperature changes alone did not determine outbreak size in 2024 and/or temperature has a non-monotonic relationship with incidence.

**Figure 2.**
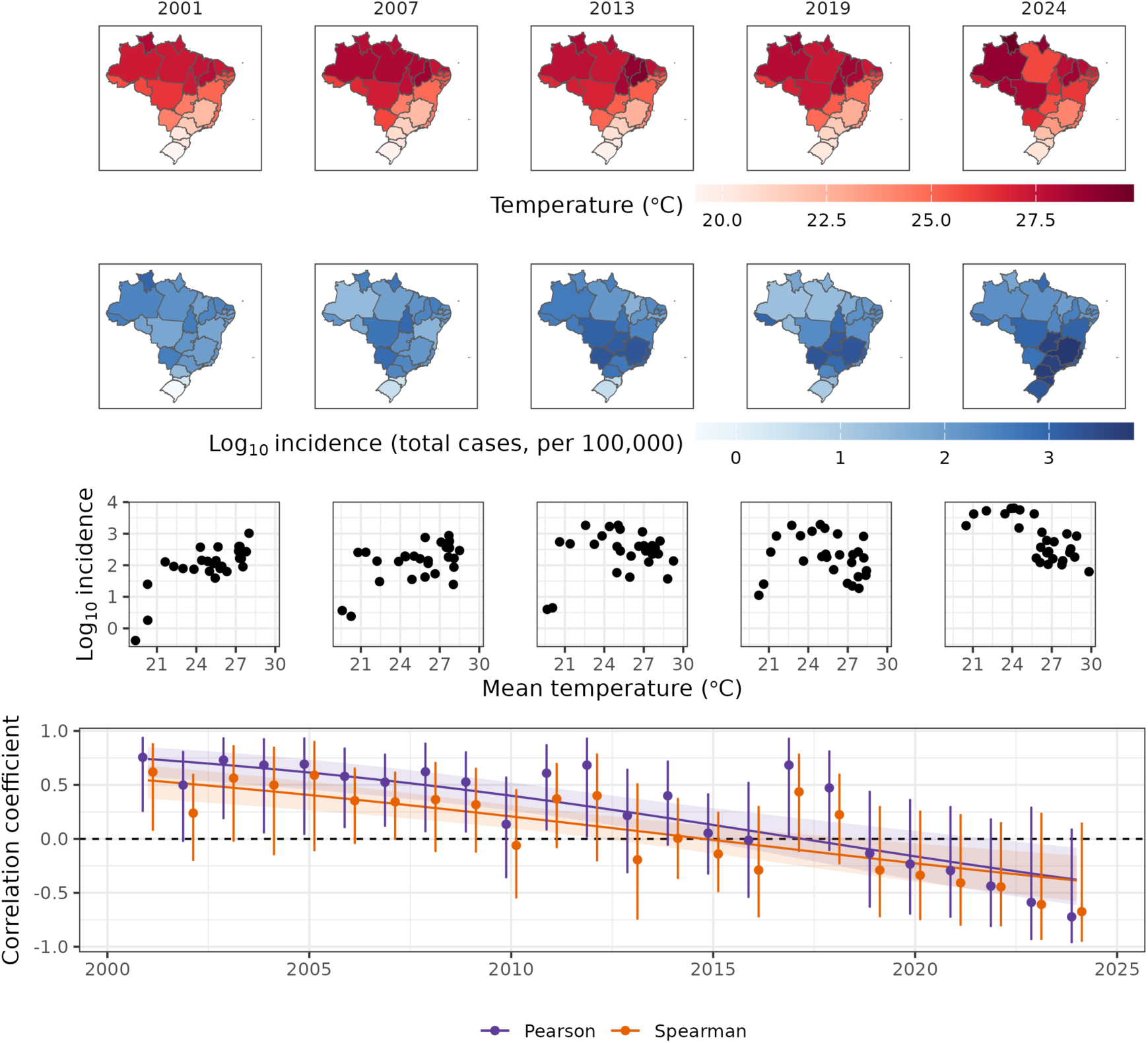
Correlations between mean temperature and incidence. Top panels (maps) show the relative spatial distributions of mean temperature (darker reds being higher), and log_10_ cumulative incidence per 100,000 (based on total cases) by state (darker blues being larger), for five example seasons. Middle panels (scatter plots) show the relationships between mean temperature and log_10_ cumulative incidence per 100,000 for the same five seasons. The bottom panel shows the Pearson and Spearman spatial correlation coefficients between mean temperature and log cumulative incidence for each season between 2001 and 2024 (points, with error bars indicating 95% confidence intervals), together with the fitted linear trends and their 95% band (Methods). Each correlation is computed across the 27 states (one point per state) within a single July-June season, between that season’s population-weighted mean temperature and its local incidence per 100,000; it is thus a spatial, cross-state correlation, and the panel shows how that spatial relationship changes from season to season. The correlation declined significantly across the series; the fitted trend is negative from the mid-2010s and its confidence band excludes zero for the most recent seasons. See SI Appendix, Fig. S2 for the equivalent figure using confirmed cases only.

### Transmission remains high at elevated temperatures, but declines with immunity

The shift in relationship between temperature and incidence, and particularly the shift towards negative correlations in recent seasons, could have been driven by temperatures in the northern states becoming too high for effective transmission^24^, or by spatial variation in immunity (higher in the north, lower in the south). Fig. 2 shows that incidence in warmer states did not noticeably drop during the 2024 season, arguing against temperature directly limiting transmission. Nevertheless, to test these hypotheses while accounting for epidemic dynamics, we estimated weekly reproductive numbers (*R_t_*) for each state using time series of weekly cases, and then fit a generalized additive model (GAM) with temperature as a smooth predictor. This approach allows us to detect whether and to what extent transmission declines at high temperatures. We found that *R_t_* increased with temperature up to 28.4°C (95% CI: 27.9 - 28.9°C; solid line in the top panel of Fig. 3), and decreased thereafter. Though *R_t_* decreased with temperatures increasing above 28.4°C, we found that they remained high (for example, *R_t_* at 32°C was higher than 70% (95% CI: 66 - 75) of observed values across the country and time). Notably, when the fitted temperature response is evaluated at each two-week window of the 2024 season and averaged across the season, its contribution to log(*R_t_*) was higher for the northern and northeastern states (+0.08 to +0.11 for Amapá, Roraima, Amazonas, Piauí, Ceará, and Maranhão) than for São Paulo (−0.09), reflecting the north’s sustained near-optimal temperatures through most of the year versus São Paulo’s briefer warm season. Temperature alone would therefore predict greater transmission potential in the north than in São Paulo, making it difficult to attribute the north’s comparatively modest 2024 incidence to thermal suppression. Note, however, that it is at the extremes where there is least data to inform the decline in *R_t_* with precision.

**Figure 3.**
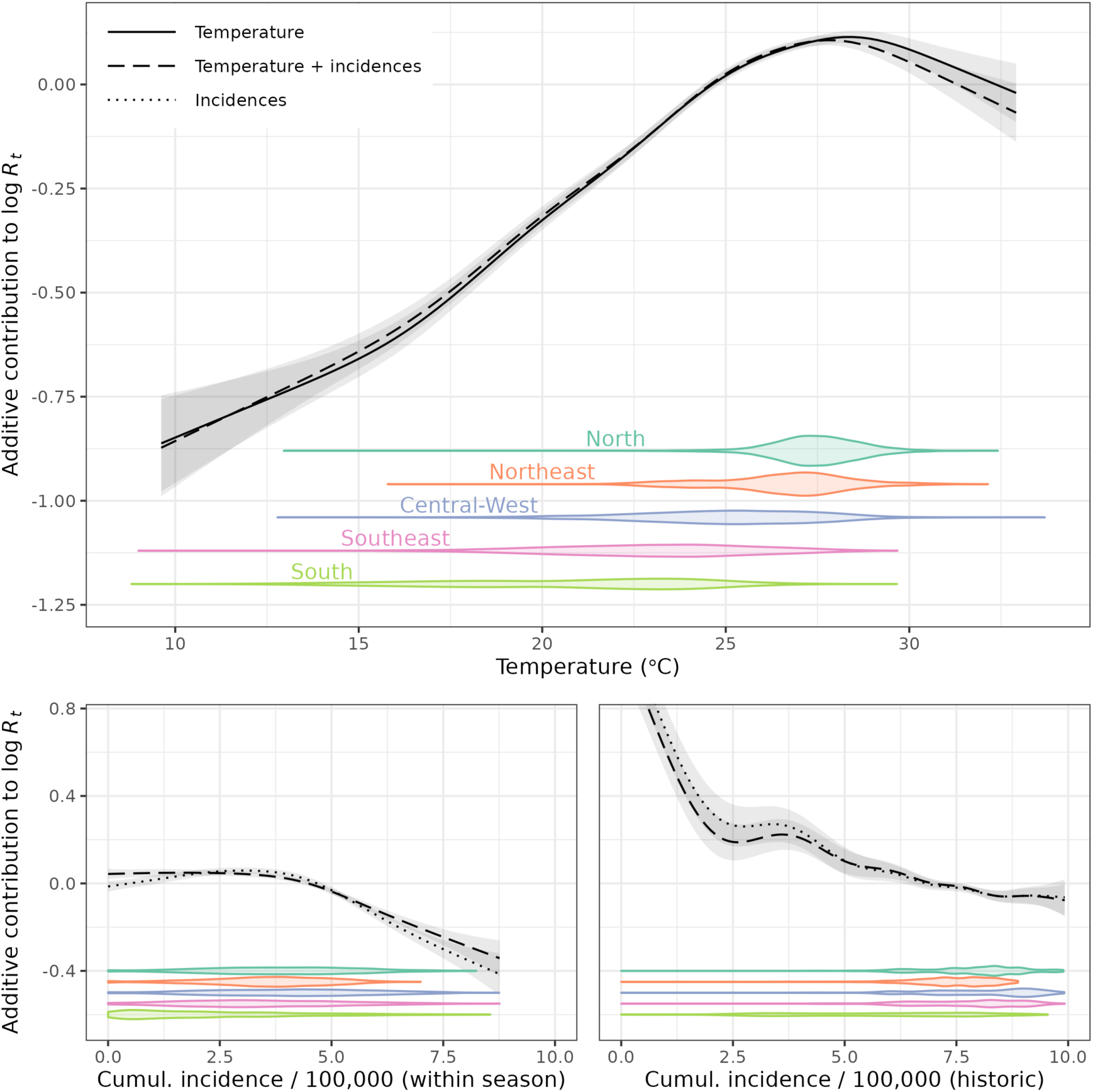
Effects of temperature and immunity on effective reproductive numbers. We fit three generalized additive models to the log-effective reproductive numbers Rt estimated using EpiEstim: (i) temperature only (top panel, solid line); (ii) temperature and lagged cumulative incidences (both within season and historic, i.e., over the whole study period, used here as proxies for susceptibility; dashed lines in all panels); and (iii) lagged cumulative incidences only (dotted lines in the bottom two panels). Violin plots at the bottom of each panel show the distributions of those variables, per region. In the temperature-only model, reproductive numbers at high temperatures remained high (70th percentile at 32℃). Instead, in the model accounting for immunity, reproductive numbers clearly decreased with increasing lagged incidences. See SI Appendix, Fig. S4 for the equivalent figure using confirmed cases only.

Given that the fitted temperature contribution to log(*R_t_*), averaged across two-week windows of the 2024 season, was higher in the northern and northeastern states listed above than in São Paulo, we next asked whether historical incidence could explain variation in transmission through the accumulation of immunity. We incorporated cumulative incidence at multiple time scales (within each season and over our entire dataset) as proxies for susceptible depletion. We found clear decreases in *R_t_* as cumulative incidence increased at both timescales, supporting a strong role for susceptibility (Fig. 3). Including these cumulative incidence terms increased the deviance explained from 12.9% to 17.4%; a model including only the cumulative incidence terms explained less deviance (6.8%) than temperature alone (12.9%), though the combined model explained more than either alone, indicating that temperature and susceptibility capture largely independent components of variation in *R_t_*. The inclusion of cumulative incidence did not appreciably change the estimated temperature response function (dashed vs. solid line in the top panel of Fig. 3): the two models’ confidence envelopes overlapped across the full range of observed temperatures. Similarly, the patterns in *R_t_* with cumulative incidences barely changed with and without the inclusion of temperature (dotted vs. dashed lines in the bottom panels of Fig. 3). The corresponding analyses using confirmed cases are shown in SI Appendix, Fig. S4.

### Changing correlations between temperature and incidence reflect spatial variation in susceptible depletion

To understand the circumstances that might give rise to reversals in the relationship between temperature and incidence, while also including the effects of immunity, we ran simulations using synthetic temperature time series with varying spatiotemporal patterns. The four scenarios (constant, linearly increasing, step increase, and spatially homogeneous high temperature; Fig. S5) are deliberate idealizations, each constructed to isolate one feature, rather than to reconstruct Brazil’s actual trajectory. These simulations show the correlation between temperature and incidence can change signs over time (SI Appendix, Fig. S6), as we observed in the data (Fig. 2). At the time of introduction, when populations are fully susceptible, the correlation starts positive. However, as susceptibles are rapidly depleted in warmer northern states but not in cooler southern states, later outbreaks shift southward, despite the lower temperatures. This causes the correlation to turn negative (the timing of the sign change depends on whether mean temperatures remain constant or increase linearly). Once susceptibles are also depleted in southern states, the correlation returns to positive. Similarly, a sharp increase in mean temperature across all states (due to, e.g., an unusually warm year) can also cause a positive correlation to temporarily turn negative (SI Appendix, Fig. S6). In contrast, removing spatial variation in temperatures and thus in immunity (assuming similarly high temperatures throughout Brazil) causes the correlation to turn negative just after dengue introduction, but it returns to positive much sooner, and remains positive throughout the simulation for most parameters explored. These simulations show that the shifting temperature-incidence correlation observed in Brazil may arise from spatial variation in immunity that results from spatial variation in temperature, rather than from temperature’s direct effect on transmission alone.

### Mechanistic estimates of susceptibility illustrate the temperature-immunity trade-off

Our GAM analysis, which revealed strong dependencies between reproductive numbers, temperature, and proxies for immunity, is a phenomenological approach that cannot distinguish how temperature affects transmission (whether directly, via effects on the thermal biology of the vector, or indirectly, through past transmission shaping immunity levels). To more explicitly estimate susceptibility and examine how it interacts with temperature in driving transmission and the role it played during the 2024 season outbreak, we estimated an initial susceptibility index (*S*^init^_index_; an estimate of the proportion susceptible at the start of a season) in each state for each season using a published temperature-dependent mechanistic SEIR-SEI model. Our estimates of *S*^init^_index_ show significant variation over time, particularly in southern states (SI Appendix, Fig. S7). Spatially, susceptibility indices were higher in the south, particularly in São Paulo, Goiás, Distrito Federal, and Minas Gerais, and lower in northern states (SI Appendix, Fig. S8). The relationship between inferred initial susceptibility and observed incidence is shown in SI Appendix, Fig. S9. While numbers cannot be directly compared, these broad patterns are consistent with regional dengue seroprevalence in participants in a vaccine trial in 16 sites across Brazil: similar seroprevalence was observed in the North, Northeast, and Central-West, with lower seroprevalence in the Southeast and substantially lower seroprevalence in the South^27^. As the correlations between mean temperature and incidence demonstrated, outbreak sizes are not solely driven by higher temperatures. In 2024, the larger outbreaks occurred in states with a relatively low mean temperature but a comparatively high susceptibility (e.g., Minas Gerais, São Paulo, and Distrito Federal; Fig. 4; SI Appendix, Fig. S10). Conversely, locations with low susceptibility required a particularly high temperature (and thus transmission) to generate a larger-than-normal outbreak (e.g., Amapá). The patterns of Fig. 4 also describe a constraint, given that high temperatures and high susceptibility rarely co-occur; sustained high temperatures deplete susceptibles, preventing this combination except transiently. Season-specific relationships are shown in SI Appendix, Fig. S11. This negative relationship between temperature and susceptibility is also consistent with theory, as seen when plotting final outbreak size as a function of temperature and susceptibility (Fig. 4 inset).

**Figure 4.**
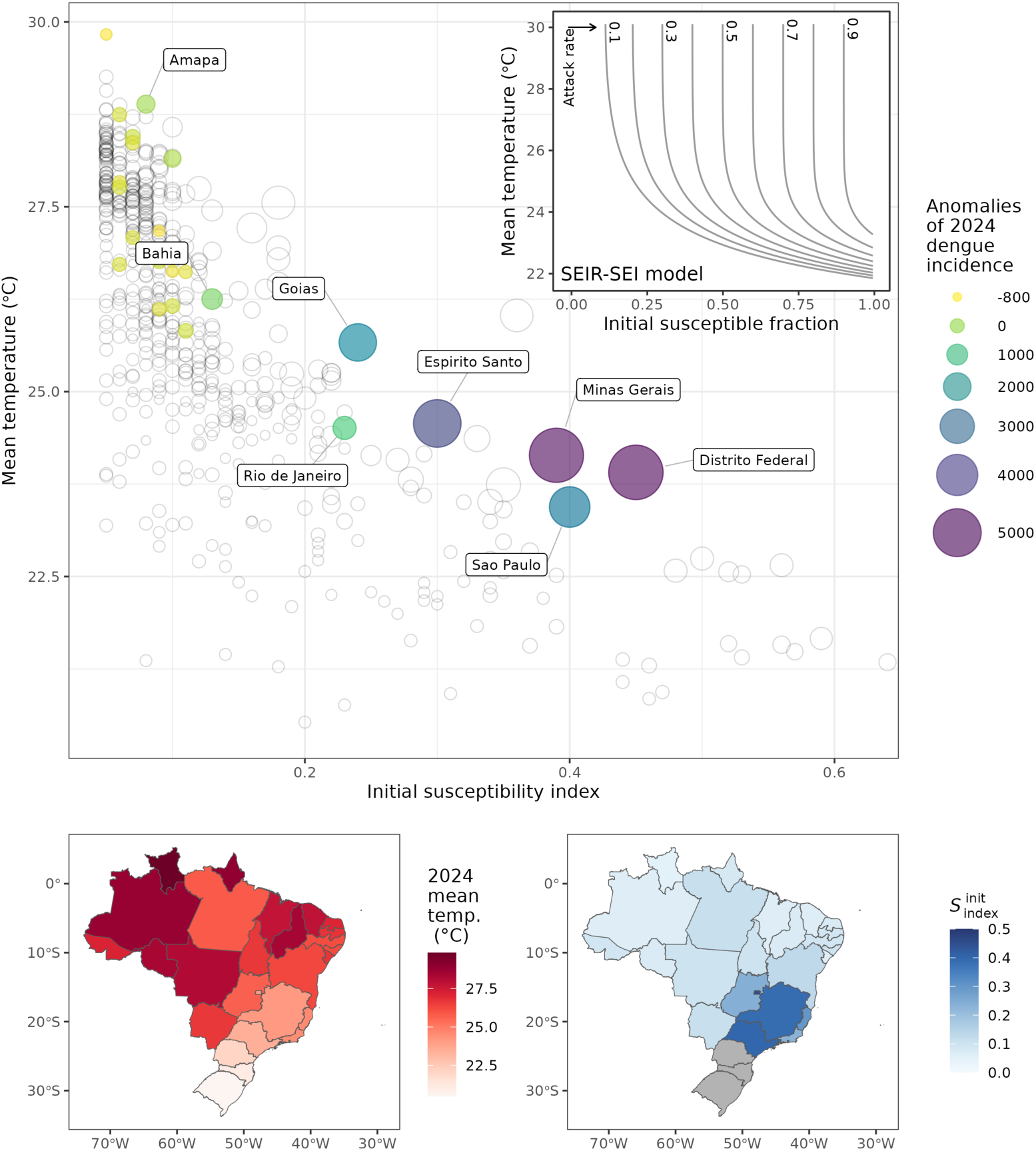
The relationship between mean temperature, the susceptibility index, and incidence anomaly. Top panel: each state’s initial susceptible population index *S*^init^_index_ and mean temperature over the 2001 to 2024 seasons. The radius of all points represents the dengue incidence (per 100,000) anomalies. The 2024 season is highlighted as filled points with colors mapping onto the incidence anomalies. States experiencing highest incidence anomalies in 2024 are labeled. The inset panel provides the theoretical relationship between initial susceptible fraction and mean temperature, with contours representing the final epidemic attack rate, as obtained from the SEIR-SEI model. This panel illustrates the negative relationship between the susceptibility indices and temperature. Bottom panels: mean temperature during the 2024 season (left) and initial susceptible proportion index *S*^init^_index_ for the start of the 2024 season (right). Note that in this analysis, the three southernmost states were excluded (shown in gray in the maps; see Methods). See SI Appendix, Fig. S10 for the equivalent figure using confirmed cases only. Mean temperature here is the season mean. It is closely correlated with peak-transmission temperature across states, so the rank order of states is largely unchanged. The two differ in scale, with strongly seasonal southern states several degrees warmer on a peak-season basis while equatorial states barely move, so a peak-season axis would span a narrower temperature range. The susceptibility indices themselves are fit to daily temperatures and are unaffected by this choice of summary.

### Both temperature and susceptibility are necessary to predict the 2024 outbreak

To assess whether temperature or susceptibility were more strongly associated with spatial variation in the 2024 outbreak, we fit three linear regression models to state-level incidence per season from 2001-2023: one using mean annual temperature, one using the lag-one final susceptibility index (*S*^final^_index_), and one including the interaction of the two. These two predictors aim to assess two different hypothesized effects of temperature on incidence: 1) mean temperature contemporaneously affects transmission, 2) accumulated immunity generated by past, temperature-driven transmission affects incidence. The second effect is not simply a lagged temperature effect, because the immunity a population carries depends on whether past outbreaks actually took off (a nonlinear function of temperature, prior susceptibility, and timing of viral introductions) and on non-thermal processes such as demographic turnover and serotype dynamics. We then assessed each model’s ability to predict observed 2024 log₁₀ incidence (Fig. 5; SI Appendix, Fig. S12). The temperature-only model performed worst, with a correlation between predicted and observed values of *r* = 0.72, a Lin’s concordance correlation coefficient *ρ*_c_ = 0.04, and a root-mean-square error RMSE = 0.75. The coefficient in the model fit was not significant, reflecting the changing relationship between temperature and incidence over the study period (Fig. 2). The *S*^final^_index_ alone performed moderately better (*r* = 0.80; *ρ*_c_ = 0.27; RMSE = 0.64 - a 15% reduction in RMSE relative to the temperature-only model), while the interaction model outperformed both (*r* = 0.82; *ρ*_c_ = 0.80; RMSE = 0.37 - a 51% and 43% reduction in RMSE over models with temperature and *S*^final^_index_ alone, respectively; Fig. 5, top panels). Both temperature-only and *S*^final^_index_-only models substantially underpredicted outbreak sizes in states reporting the largest 2024 outbreaks, whereas the interaction model was able to better capture this variation (Fig. 5, bottom panels), though some states, notably Espírito Santo, were still systematically under-predicted by the interaction model (Fig. 5). The interaction coefficient for the model fit to 2001-2023 was strongly significant (*β* = 1.44, *p* = 1.01 x10^-14^; although note this *p*-value accounts for neither spatial autocorrelation nor the estimation uncertainty in *S*^final^_index_, which is a model-derived rather than observed covariate whose estimates for a state agree only weakly from one year to the next; Fig. S13), showing that temperature’s effect on incidence increases with higher susceptibility, and likewise, susceptibility’s effect is greater at higher temperatures. We also compared this linear interaction model against a non-linear (GAM) version of the same model. Although the non-linear fit captured genuine curvature within the training data, it predicted the out-of-sample 2024 season less well, supporting our use of the simpler linear formulation.

**Figure 5.**
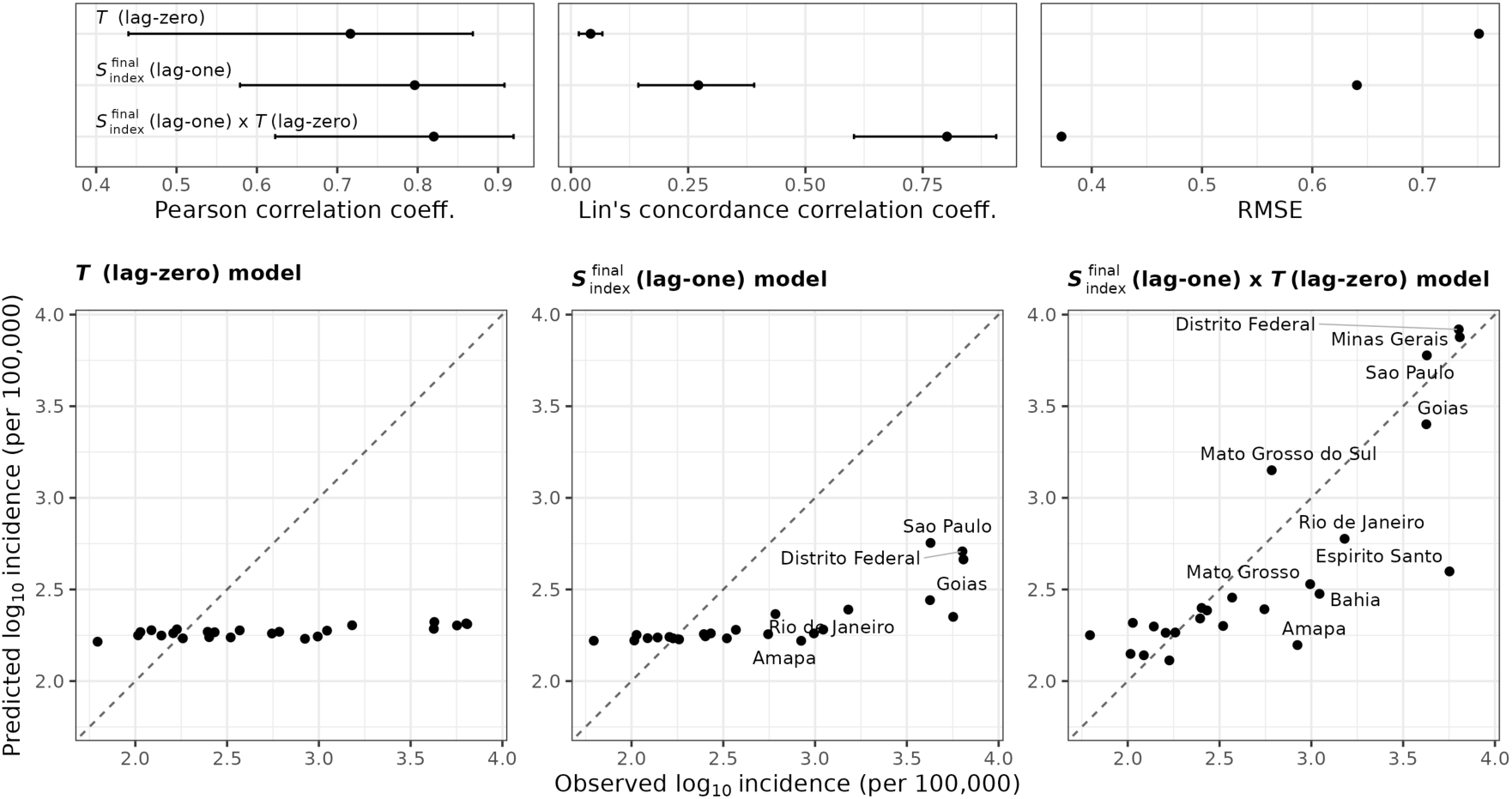
Predicting the 2024 outbreak with temperature and susceptibility. Performance of three linear regression models assessed by Pearson correlation coefficients, Lin’s concordance correlation coefficients and RMSEs, between predicted and observed 2024 season log₁₀ incidence (top panels). Bottom panels show predicted vs. observed values for the three models: using temperature only (left), final susceptibility index (lag-one *S*^final^_index_) only (middle), and their interaction (right). The model including both susceptibility and temperature clearly outperformed models only including one of the two covariates. See SI Appendix, Fig. S12 for the equivalent figure using confirmed cases only.

## Discussion

Brazil experienced an unprecedented dengue season in 2024, coinciding with record high mean temperatures across much of the country. This prompted climate change to be proposed as a primary cause, yet the largest outbreaks occurred in southeastern states with comparatively lower temperatures than northern states (Fig. 1), culminating in the shift to negative spatial correlations (Fig. 2). We show this apparent contradiction is explained by spatial variation in population immunity: the 2024 surge resulted from an increasingly suitable climate interacting with areas of high susceptibility. Significant clusters of incidence anomalies aligned with states we inferred as having relatively high susceptibility (e.g., São Paulo, Minas Gerais, Distrito Federal), while states in the North and Northeast, which also experienced warming, had lower susceptibility and reported more limited outbreaks (SI Appendix, Fig. S8). We hypothesized this pattern could arise either from temperatures in northern states becoming too high for efficient transmission, or from geographic variability in susceptibility. Analysis of effective reproductive numbers supported the latter hypothesis: transmission at high temperatures remained substantial (70th percentile of the *R_t_*-temperature curve), while models incorporating susceptibility proxies explained significantly more variation and showed clear declines in *R_t_* with cumulative incidences (Fig. 3). Finally, we used a mechanistic SEIR-SEI model to estimate a susceptibility index for each state and season, quantifying the spatial variation in immunity (Fig. 4). Out-of-sample predictions for 2024 showed that both temperature and the susceptibility index were necessary to adequately predict the state-level 2024 incidences (while susceptibility and temperature, in isolation, performed comparatively poorly; Fig. 5). The significant interaction between temperature and susceptibility suggests that temperature’s association with incidence is modulated by susceptibility: the effect of temperature is greater with high levels of susceptibility, and similarly, the effect of susceptibility on incidence is greater at higher temperatures.

The observed shift from positive to negative fitted correlations between incidence and temperature could serve as a signature of range expansion into immunologically naive populations. The relationship between dengue incidence and temperature is mediated by immunity, and can be expected to evolve, from when a virus is initially introduced, to endemicity. In the initial phases post-introduction to climatically suitable areas and immunologically naive populations, a positive correlation between incidence and temperature might be expected, though some variability may arise due to differences in the timing of introduction of DENV to new areas. However, as susceptible hosts are depleted in these optimal environments, the spatial patterns of incidence would change. Climate-driven expansion of the vector’s range into new, immunologically naive areas, particularly large metropolitan centers, could compound this effect. Together, these processes would lead to an increasingly heterogeneous immune landscape where low susceptibility blunts outbreaks despite more optimal temperatures, resulting in the observed negative correlations. In the future, as dengue becomes endemic and Brazil becomes more immunologically homogeneous, we might expect the correlation between temperature and incidence to become positive again.

The role of population immunity in explaining dengue outbreaks is often overlooked, but crucial given the role played by range expansion in the 2024 outbreak in Brazil. In a recent analysis of 21 mostly dengue-endemic countries, using case data up to 2019, Childs et al. (2025)^7^ estimated the average temperature-incidence relationship across locations and tested whether it varies with population immunity, using lagged incidence as a proxy. Finding it similar across immunity levels, they applied a single temperature response when projecting the effects of warming. Our *R_t_* analysis is broadly consistent: the temperature response is similar with and without the cumulative-incidence terms (top panel of Fig. 3). That the response is largely immunity-invariant does not, however, mean immunity is unimportant for outbreak magnitude. The cumulative-incidence terms explain significant variation in *R_t_*, and between states it is the susceptibility index that separates those that surged in 2024 from those that did not (Figs. 4 and 5). The results are compatible because they answer different questions. Childs et al. estimate an effect averaged across many endemic locations and years, not a single year or spatial anomaly, and attribute the immunity-invariance to endemic populations never reaching persistent herd immunity, through waning and serotype turnover. The 2024 outbreak in Brazil points to the opposite case: large populations with lower historical transmission undergoing a range expansion. This is a scenario their analysis explicitly sets aside. Consistent with this, our analyses suggest that accounting for immunity greatly improves predictions of the spatial patterns of the 2024 outbreak

Defining *R*_0_, and especially the conditions that define the critical transmission threshold *R*_0_ = 1, remains critical for predicting dengue incidence. However, mechanistic expressions for the temperature dependence of *R*_0_ usually assume stationarity^28^, and rely on variables that are difficult to estimate in the field and are expected to be spatially heterogeneous, such as the vector-to-host ratio (on which *R*_0_ depends; see SI Appendix, Appendix S2). Studies deriving and applying temperature-dependent *R*_0_ models have therefore focused mainly on the shape of its temperature dependence, the transmissive range (*R*_0_ > 0), or the temperature that maximizes *R*_0_, rather than on absolute *R*_0_ values^3,6^. This limitation bears directly on why the Southeast surged in 2024 and not earlier: without an absolute estimate of *R*_0_, we cannot identify whether or when its transmission potential first crossed the *R*_0_ = 1 threshold for sustained transmission, and so cannot fully account for the timing of the outbreak. It is also possible that preceding years saw widespread but largely unreported transmission (primary dengue infections are frequently mild or asymptomatic) with part of the 2024 reported surge reflecting secondary, more often symptomatic infections in populations primed in the year(s) before.

Our findings are subject to several limitations. Surveillance biases likely distorted incidence data: the misclassification of Oropouche virus (OROV) or Chikungunya cases as dengue in the South (e.g., Santa Catarina) may have inflated reported incidence, while systemic underreporting of DENV-3 in the Northeast may have masked earlier resurgences, thereby skewing our susceptibility estimates for these regions^9,15^. Interaction with health care resources or attention may result in time- and incidence-varying reporting rates of dengue (e.g., large outbreaks may increase reporting due to broader recognition of dengue risk or decrease reporting due to saturation of reporting resources). Our estimates of reproductive numbers assume a fixed generation interval, which instead would be expected to vary with temperature^29,30^. Our modeling framework makes several simplifying assumptions. The single-serotype SEIR-SEI model does not account for serotype-specific immunity; our ‘initial susceptibility index’ is therefore an aggregate measure that may mask complex inter-serotype immune dynamics. Because each state-season is fit independently, with no constraint linking successive seasons, a serotype introduction that reset susceptibility would be absorbed as a jump in *S*^init^_index_; the fit is agnostic as to whether a given shift reflects a serotype introduction, demographic turnover, or waning cross-protection.

We have not addressed several alternative mechanisms that could have contributed to the pattern of dengue observed in 2024 in Brazil. Introduction of new serotypes into some areas may have contributed to greater levels of incidence. State-level serotype composition for 2024 and regional serotype composition for 2007–2024 among typed surveillance records are shown in SI Appendix, Figs. S14 and S15, respectively. These observations caution against assuming uniform serotype circulation across Brazil, but do not support large spatial differences across Brazil in serotypes circulating. The contribution of pre-existing immunity to higher rates of apparent illness was not explicitly addressed here, though could also contribute to variation. Finally, differences in reporting, though estimated in our models and not found to contribute to systematic differences in cases in southern states compared to northern states, could contribute to differences in incidence.

All analyses were conducted at the state level. We chose this scale because over a third of Brazil’s municipalities have a median of zero cases per season (five or fewer, for almost two-thirds), and 39% of all municipality-seasons record zero cases, a degree of sparsity that destabilizes both reproductive-number estimation and susceptibility inference. Confounding requires that an omitted factor be correlated with temperature: sub-state heterogeneity in incidence that is uncorrelated with temperature cannot bias the state-level association, and averaging over many such independent local departures suppresses their contribution to the state-level estimate. The risk aggregation cannot address is heterogeneity that is correlated with temperature at the sub-state level, e.g., municipality-level gradients in reporting rate or vector-control investment that correlate with local climate, which could bias the state-level associations in either direction. The *R*_t_ GAMs include state and season fixed effects, which absorb time-invariant between-state differences and nationwide inter-seasonal variation, but not state-specific time-varying confounds or sub-state heterogeneity. The correlation analysis (Fig. 2) and the regression models (Fig. 5) make no such adjustment, because the between-state contrast is itself the quantity of interest. We therefore cannot rule out residual confounding at the state level. Moving to a municipality scale would not in itself resolve this: separating such confounding from the associations of interest would still require either measured covariates for the suspected confounds (health-system capacity, vector-control measures, reporting rates), or a design that removes variation at the confound’s own scale (e.g., within-municipality, over time), which reintroduces the data sparsity that motivated the state-level choice in the first place. In inferring the initial susceptibility index, we also assumed a spatiotemporally uniform vector-to-host ratio, reporting rate, and number of infections introduced to each location at the start of a season, which are significant oversimplifications. Spatial differences in vector control act through the vector-to-host ratio: historically low-burden states such as São Paulo may never have built the dengue-specific control capacity of more affected states, and any such difference would be absorbed into the susceptibility index, producing a spatial pattern much like the one we attribute to immunity. We therefore treat the susceptibility index as a relative rather than absolute measure of population susceptibility, a use supported by the stability of indices within and across states when estimated from total versus confirmed cases, despite their substantially different reporting rates (SI Appendix, Fig. S7). The assumptions made in these compartmental models also lead to unrealistically fast dynamics, potentially inconsistent with spatial heterogeneity within states, making the estimation of susceptibility more difficult. Finally, beyond the inherent assumptions of our SEIR-SEI model, we highlight three potential temperature-related biases. First, the temperature dependence of vector life-history traits, derived from laboratory experiments under constant conditions, may bias rate estimates in fluctuating real-world temperatures, with the expected bias dependent on the response functions’ curvature at a given temperature^31^. Second, the gridded temperature dataset we use here is interpolated and averaged over relatively large spatiotemporal scales, which may not reflect the microclimatic temperatures actually experienced by mosquitoes, nor the precise temperatures in the laboratory experiments used to parameterize the model. Third, as demonstrated by Childs et al.^7^, temperature datasets can also exhibit significant discrepancies for a given location, potentially leading to substantial variations in estimated vital rates solely based on the dataset used.

We used total reported dengue cases in our analyses, which may introduce biases due to misclassification of other febrile illnesses. However, when we repeated analyses using only confirmed cases, results remained qualitatively unchanged (see SI Appendix, Appendix S3). Confirmed cases have their own limitations: there can be less testing during large epidemics due to resource constraints, potentially leading to greater underreporting in high-incidence years and states. Thus, while neither measure is perfect, the consistency of results across both definitions strengthens confidence that the spatial and temporal patterns we identify, particularly with regards to the roles of temperature and immunity, likely reflect true epidemiological dynamics rather than surveillance biases.

As noted above, we do not distinguish between primary and secondary infections, limiting our ability to project past 2024. The 2024 outbreak in southeastern states could constrain transmission through accumulated immunity. However, populations with monotypic immunity remain vulnerable to secondary infections, which could lead to increased cases if other serotypes were to circulate in the near future. At the same time, continued range expansion into southern states (Paraná, Santa Catarina, and Rio Grande do Sul), and potentially other large, immunologically naive metropolitan areas like Buenos Aires, could drive substantial future outbreaks.

Our results suggest avenues for further research to improve our understanding of the spatiotemporal evolution of dengue epidemics in emerging and re-emerging settings. First, reconstruction of population susceptibility even at crude scales (e.g., statewide and annual) will provide information about areas at risk of increased transmission and with higher sensitivity to climatic changes. This reconstruction could be achieved using case data where available^32^, bolstered by population representative serological surveys^27,33^, although methodological development is necessary to integrate data sources and project these estimates countrywide. Second, our models using reconstructed susceptibility indices outperformed models using mean temperature, suggesting that current models using temperature-dependent reproductive numbers to predict changes in dengue transmission would also benefit from integration of susceptibility estimates informed by transmission models. One step towards this could be the use of effective reproductive numbers in assessing climate associations (rather than incidence) as this accounts, at least in part, for changes in susceptibility.

Consideration of population immunity will likely also be critical in improving the accuracy and increasing the time horizon of models forecasting dengue burden, as noted by Wu et al.^34^. The majority of published forecasting models use only climate data as predictors^35^; evaluation of models on Brazil’s 2024 epidemic highlights the limitations of such models, as observed by Araujo et al.^36^. Whether a measure of immunity is needed for prediction in other settings will depend on regime: it is likely to matter most during susceptibility regime shifts such as range expansions into newly suitable areas, or in the aftermath of a serotype introduction or unusually large outbreaks, and least where susceptibility is more stable, as in long-established endemic areas. Our results also note the weakness of using only historical case numbers to project future burden. Integration of historical climate and estimates of immunity (including serotype specific estimates) into forecast models would represent significant progress in this field and increase the value of these models to public health.

## Materials and Methods

### Study design

We conducted a retrospective ecological time-series analysis to investigate the drivers of the 2024 dengue epidemic in Brazil. Our analysis compared state-level incidence, temperature, and their 2024 season anomalies against a 23-year historical baseline (2001–2023).

### Data collection and processing

We obtained and processed individual-level dengue case reports from Brazil’s SINAN (Sistema de Informação de Agravos de Notificação) database (2000–2024) using the microdatasus R package ^37^. We retained confirmed or suspected cases, excluding implausible records (e.g., invalid ages). All analyses were performed on total cases (both confirmed and suspected); we repeated analyses using confirmed cases only, and provide these results in the SI. Where the symptom onset date was later than the notification date (n = 25), we used the notification date as a proxy for symptom onset.

Municipality and state identifiers were standardized with Instituto Brasileiro de Geografia e Estatística (IBGE) codes, prioritizing the residence municipality when the notification location was missing. We aggregated daily state-level counts of suspected, confirmed, and fatal cases into time series for the period of July 1, 2000, to June 30, 2024. A July-to-June epidemic year was defined to align with dengue seasonality (where, e.g., the 2024 season refers to July 2023-June 2024). Unless otherwise specified, references to a year or season follow this definition. Daily incidence rates per 100,000 were calculated using IBGE population estimates, and we calculated annual incidence as the total number of reported cases in a season per 100,000. We conducted analyses on 27 Brazilian states.

For supplementary analyses, we summarized DENV-1–4 composition among surveillance records with a valid serotype assignment by state for 2024 (SI Appendix, Fig. S14) and by region for 2007–2024 (SI Appendix, Fig. S15), using source-file years rather than epidemic seasons. Serotype information was unavailable in the SINAN data for 2000–2006. We displayed compositions only for states with at least 200 typed records and region-year combinations with at least 50 typed records; these proportions do not estimate population-level serotype prevalence.

Daily gridded minimum and maximum temperatures were downloaded from NOAA (https://psl.noaa.gov/data/gridded/data.cpc.globaltemp.html; February 2025), and gridded population data was downloaded from SEDAC (https://sedac.ciesin.columbia.edu/data/set/gpw-v4-population-count-rev11/data-download; June 2022). The daily mean temperature for each grid cell was first calculated as the mean of the minimum and maximum temperatures. For each state, a population-weighted mean of these daily mean temperatures was then calculated to best estimate the temperature affecting human-mosquito populations and interactions.

### Descriptive and statistical analyses

To quantify the relationship between temperature and incidence, we calculated Pearson’s correlation coefficients (*r*) and associated 95% confidence intervals, across states (one value per state), between (i) the 2024 season temperature anomalies and the 2024 incidence anomalies, and (ii) season-mean temperature and log_10_ season-total cumulative incidence per 100,000, computed separately for each season from 2001 to 2024. For (ii) we also calculated the Spearman rank correlation, as a robustness check insensitive to the shape of the temperature-incidence relationship. Anomalies are defined per state as the 2024-season value minus the mean of the 2001-2023 season values, using season-aggregated quantities, i.e., one anomaly per state. To account for spatial autocorrelation, we adjusted the effective degrees of freedom using Dutilleul’s^38^ modified *t*-test (R package SpatialPack^39^), with pairwise distances computed between population-weighted state centroids projected to EPSG:5880 (SIRGAS 2000 / Brazil Polyconic). Spatial analyses were conducted using the spdep^40^ v1.3-11 R package.

To summarize how the season-by-season correlation series changed over time, we regressed the coefficients on season by generalized least squares on the Fisher-z scale (atanh *r*), with inverse-variance weights and a first-order autoregressive term across seasons, to account for the coefficients’ unequal uncertainty and serially correlated residuals. Trend slopes are reported as the change in atanh *r* per year; fitted values are back-transformed to the correlation scale for Fig. 2.

### Modeling framework

#### Variation in reproductive numbers with temperature and immunity

To understand how spatiotemporal patterns in dengue cases in Brazil are related to temperature and population susceptibility, we first analyzed an outcome of estimated effective reproductive numbers (*R_t_*) derived from reported case data. *R_t_* are more robust to differences in reporting than raw case numbers, and reflect a combination of transmissibility and population immunity (*R_t_*=*R*_0_\**S_t_*, where *S_t_* is the proportion of the population susceptible). Reproductive numbers were calculated using the EpiEstim v2.2.5^41^ package in R from weekly state-level time series of cases over a two-week moving window, assuming a mean serial interval of 12 days and a standard deviation of 3 days. Estimates based on fewer than five cases within a window were deemed unreliable and excluded from subsequent analysis, and *R_t_* values equal to or greater than 10 were also deemed unreliable and excluded (as they were often estimated for the start of the time period, or in states that reported few cases). The log-transformed *R_t_* values were the dependent variable in a generalized additive model (GAM) from the mgcv v1.9.4 package^42^ in R. The GAM included a smooth term for mean temperature for the same two-week moving window, and smooth terms for the lag-1 log-cumulative incidence per 100,000 (both within-season and overall, state-level) as a proxy for the build-up of immunity prior to the time window over which *R_t_* was estimated, and thus accounting for changing susceptibility driven by annual outbreak dynamics and historical transmission. We used a contemporaneous temperature window rather than distributed lags deliberately: the aim is to isolate temperature’s direct effect on transmission from its lagged, indirect effect through accumulated immunity, which the cumulative-incidence terms (themselves a historical function of temperature) already capture. Adding lagged temperature would induce collinearity between it and the lagged incidences, blurring this decomposition. More fundamentally, a temperature lag cannot represent the path-dependent depletion of susceptibles - whether an outbreak actually occurred - that the cumulative incidence terms carry. We note that “direct” is approximate here, since temperatures ultimately act on mosquito life history at multiple lags. Season and state were included as categorical fixed effects. To assess the relationship between temperature and *R_t_* when not accounting for historic incidence, the same model was fit excluding the cumulative incidence smooth terms. To minimize the risk of overfitting, the GAM was fitted with double penalization and the smoothing parameter estimation was made using restricted maximum likelihood. To further reduce the risk of overfitting, we performed 10-fold cross-validation to select the optimal basis dimension (*k*) from a range of 3 to 30 (the same value used across all smooth terms). To account for temporal autocorrelation in the residuals, models were fitted using the ‘bam’ function with an AR(1) error structure. The autocorrelation parameter *ρ* was estimated from the mean lag-1 autocorrelation of residuals from an initial model fit, calculated separately for each state-year series. Performance was measured by the RMSE and Pearson correlation coefficient between predicted and observed values on the out-of-sample data.

#### Characterizing susceptibility with a temperature-dependent single-serotype SEIR-SEI model

Next, to test more precise hypotheses of the role of susceptibility and temperature, we fit a mechanistic temperature-dependent dengue transmission model^43^ to the same data from Brazil, and used it to estimate susceptibility. We implemented a two-step procedure: first, we set up a compartmental model with susceptible, exposed, infectious, and removed humans (SEIR) and susceptible, exposed, and infectious mosquitoes (SEI). Temperature dependence was incorporated through mechanistic parameters (mosquito biting rate, infection probability, etc.) with functional relationships validated by laboratory experiments (Mordecai et al., 2017)^3^. Using daily temperature as input, we numerically solved the system of ordinary differential equations to derive dengue incidence time series for each state and season, estimating the cumulative incidence as the number of cases transitioning to the recovered (*R*) compartment (see SI Appendix, Section S1 for model formulation and Table S1 for parameters). Second, we performed a grid search over four parameters (ratio of vectors to humans *M*, initial infected proportion *I_h_*, reporting rate *a*, and initial susceptible proportion *S*^init^_index_; see SI Appendix, Table S2 for the values used) to identify the susceptible fraction for which the model predictions best matched observed cases in each state and season. For each parameter combination, state, and season, we generated predicted incidence using population-weighted temperature data and calculated the Poisson negative log-likelihood (NLL) against observed monthly cases. We extracted the *S*^init^_index_ minimizing NLL for each state and season and parameter combination, then summed NLLs across all states and seasons to find the optimal values of *M*, *I*_h_, and *a*, producing time series of *S*^init^_index_ for each state.

Finally, with these *S*^init^_index_ and best-fit parameter set, we simulated the number of cases in each state for each season, and extracted the final susceptible proportion *S*^final^_index_, the proportion of the population still susceptible at the end of the season. We compare the *S*^final^_index_ for year *y* - 1 with the *S*^init^_index_ for year *y* in SI Appendix, Fig. S13. Due to limited incidence in the three southernmost states (Rio Grande do Sul, Santa Catarina, and Paraná), these states were excluded from this analysis.

In our estimation, we acknowledge that multiple other parameters, namely *M*, *I_h_*, and *a,* could vary across states and over time, and so the set of *S*^init^_index_ we find likely absorbs some of the spatiotemporal variation in those parameters. Thus, we interpret the *S*^init^_index_ not as absolute estimates of starting susceptible proportions but a proxy that is correlated with the actual susceptible proportions.

### Simulations to study relationships between temperature and incidence over the transition from emergence to endemicity

To explore the conditions under which correlations between temperature and incidence (estimated in the Descriptive/statistical analyses section above) might change over time, we used the same SEIR-SEI model described above, here including human birth and death rates. The observed temperature inputs were replaced with sine curves parameterized for each state to remove data-specific confounding effects and noise, and to focus on the effects of specific changes in temperature on the resulting correlations. The scenarios are not calibrated to Brazil’s observed warming; they are idealized to isolate individual mechanisms. The simulations were run for a period of 50 years. Four distinct temperature scenarios were modeled (see SI Appendix, Fig. S5):

- “Constant temperature” scenario: the mean temperature for each state was held at its observed value, while the amplitude of the sine curve was set to roughly match the observed seasonal temperature variation (see SI Appendix, Fig. S16).
- “Linearly-increasing temperature” scenario: the mean temperature was set to decrease linearly from year 50 back to year 0 at a rate of 0.73 degrees per decade.
- “Step-increase in temperature” scenario: the mean temperature was held constant (as in the “constant” scenario) until year 40, at which point the mean was increased by one degree for the remainder of the simulation.
- “Constant high temperature” scenario aimed to remove spatial variation in temperature (for instance, simulating conditions in a location where the disease is endemic). The mean and amplitude of temperature for each state was sampled from narrow normal distributions (for the mean, *μ* = 27℃, *σ* = 0.5℃; for the amplitude *μ* = 3℃, *σ* = 0.1℃), approximately corresponding to the mean and amplitude of the temperatures observed in the warmest states (SI Appendix, Fig. S16).

### The relative contributions of temperature and incidence in explaining the 2024 outbreak

We fit three linear regression models to state-level annual incidence data from 2001-2023, and assessed their ability to predict 2024 season log_10_ incidence. The models were (i) lag-zero mean temperature during the season (representing the direct effects of temperature on transmission), (ii) lag-one final susceptibility index *S*^final^_index_ (representing immunity’s role in blunting outbreaks), and (iii) the interaction between mean temperature and final susceptibility index. Model performance was assessed using the Pearson correlation coefficient, Lin’s concordance correlation coefficient, and root-mean-square error (RMSE) between predicted and observed 2024 season log_10_ incidences. All processing and analyses were conducted in R (v4.3.1).

## Data Availability

Dengue surveillance data were obtained from Brazil SINAN/DATASUS system. Temperature data were obtained from NOAAs Climate Prediction Center global temperature dataset, gridded population data from SEDAC Gridded Population of the World, Version 4 dataset, and population estimates from IBGE. Derived aggregate data supporting the analyses are available from the corresponding authors upon reasonable request, subject to applicable source-data access and redistribution conditions.

https://psl.noaa.gov/data/gridded/data.cpc.globaltemp.html

https://sedac.ciesin.columbia.edu/data/set/gpw-v4-population-count-rev11/data-download

## Acknowledgments

M.L.N. is supported by the Fundação de Amparo à Pesquisa do Estado de São Paulo (FAPESP) (grant 2022/03645-1), the Instituto Nacional de Ciência e Tecnologia em Vigilância Genômica de Vírus e One Health (INCT Viral Genomic Surveillance and One Health) (grant 405786/2022-0), and the Centers for Research in Emerging Infectious Diseases (CREID) through the Coordinating Research on Emerging Arboviral Threats Encompassing the Neotropics (CREATE-NEO) program (grant U01AI151807), funded by the National Institutes of Health (NIH, USA). M.L.N. is also a CNPq Research Fellow.

## Author Contributions

Bachir Assao Neino (Data curation, Formal analysis, Investigation, Methodology, Visualization, Writing—original draft, Writing—review & editing), Bernardo García-Carreras (Data curation, Formal analysis, Investigation, Methodology, Visualization, Writing—original draft, Writing—review & editing), Mondal Hasan Zahid (Investigation, Methodology, Writing – review & editing), Angkana T. Huang (Methodology, Writing – review & editing), Youngsuk Ko (Writing – review & editing), Paloma Cárcamo (Writing – review & editing), Luciano A. Moreira (Writing – review & editing), Mauricio L. Nogueira (Writing – review & editing), Mauro M. Teixeira (Writing – review & editing), Albert I. Ko (Writing – review & editing), Matt D.T. Hitchings (Conceptualization, Methodology, Supervision, Writing—original draft, Writing—review & editing), Derek A.T. Cummings (Conceptualization, Methodology, Resources, Writing—original draft, Writing—review & editing)

## Classification

Paste the major and minor classification here. Dual classifications are permitted but cannot be within the same major classification.

## Ethics/declarations

This study used de-identified secondary surveillance data and reports aggregate results. No identifiable participant information is included in the manuscript or supplementary materials.

