## Supplementary Information for "The role of climate and immunity in Brazil’s unprecedented outbreak of dengue in 2024"

*Corresponding.

**This PDF file includes:**

Supporting text
Figures S1 to S16
Tables S1 to S4
SI References

**Supporting Information Text**

**Appendix S1. Temperature-dependent SEIR-SEI single serotype model**

We use an SEIR-SEI model based on that of Huber *et al.* (2018)[1]. The rates of change of the human compartments are:

$$\frac{dS_h}{dT} = r_h N_h - (b(T)\, p_s(T)\, \frac{I_v}{N_h} + \mu_h)\, S_h,$$

$$\frac{dE_h}{dT} = b(T)\, p_s(T)\, \frac{I_v}{N_h} S_h - (\delta_h + \mu_h)\, E_h\ ,$$

$$\frac{dI_h}{dT} = \delta_h\, E_h - (\gamma_h + \mu_h)\, I_h,$$

$$\frac{dR_h}{dT} = \gamma_h\, I_h - \mu_h\, R_h\ ,$$

where $T$ is temperature, $N$, $S$, $E$, $I$, and $R$ are the total, susceptible, exposed, infected, and recovered number of individuals, with subscripts $h$ and $v$ denoting human and vectors, $b$ is the vector biting rate, $p_s$ is the probability of mosquito infectiousness (probability of human infection per bite of an infectious mosquito), $\delta_h$ is the intrinsic incubation period, and $\gamma_h$ is the rate of human recovery from infectiousness, and $r_h$ and $\mu_h$ are the human birth and mortality rates (see Table S1). The rates of change of the vector compartments are:

$$\frac{dS_v}{dT} = \frac{g(T)\, s(T)\, d(T)\, N_v\, (1 - \frac{N_V}{K(T)})}{\mu_v(T)} - (b(T)\ p_c(T)\, \frac{I_h}{N_h} + \mu_v(T))\, S_v,$$

$$\frac{dE_v}{dT} = b(T)\ p_c(T)\, \frac{I_h}{N_h}\, S_v - (\delta_v(T) + \mu_v(T))\, E_v\ ,$$

$$\frac{dI_v}{dT} = \delta_v(T)\, E_v - \mu_v(T)\, I_v + \varepsilon\, N_h,$$

where $g$ is the number of eggs laid per female, $s$ is the probability of egg-to-adult survival, $d$ is the mosquito egg-to-adult development rate, $\mu_v$ is the vector mortality rate, $p_c$ is the probability of mosquito infection per bite on an infectious human, $\delta_v$ is the virus extrinsic incubation rate, and with $\varepsilon$, we introduce vectors at a constant rate as a proportion of the total human population. The reason for introducing vectors is to prevent the vector population from going extinct during the colder days, given simulations are started in July (low-season in many states in Brazil, particularly in the South), and the likelihood of importations being correlated to human population size.

The vector carrying capacity $K$ is given by

$$K(T) = \frac{g(T)\, s(T)\, d(T)\, \mu_v(T)^{-1} - \mu_v(T_0)}{g(T_0)\, s(T_0)\, d(T_0)\ \mu_v(T)^{-1}}\, N_v{}^{max}\ exp\,(\frac{-E_a\,(T - T_0)^2}{k\,(T + 273.15)\,(T_0 + 273.15)})\ ,$$

where $N_v{}^{\max}$ is the maximum carrying capacity (capped at some multiple $M$ of the total human population), $k$ is the Boltzmann-Arrhenius constant, $T_0$ is the reference temperature at which carrying capacity is greatest (here, following Huber *et al.* (2018)[1], and $E_a$ is the "activation energy", that defines the rate of decline in the carrying capacity either side of $T_0$. We estimated cumulative incidence by tracking the number of cases transitioning to the recovered ($R$) compartment. See Table S1 in SI for the parameter definitions and values used.

**Table S1:** Variables used in the SEIR-SEI model, with definitions, units, and values used. Temperature-dependent rates are given by functions defined by multiple parameters. The Brière function *is* $cT(T - T_{\min})(T_{\max} - T)^{1/2}$, *and the quadratic function is* $c(T - T_{\max})(T - T_{\min})$. All values referring to mosquito rates taken from Mordecai *et al.* (2017)[2] and Huber *et al.* (2018)[1] correspond to those for *Aedes aegypti*.

| ***Symbol*** | ***Definition*** | ***Units*** | ***Function type*** | ***Value(s)*** | ***Ref.*** |
|---|---|---|---|---|---|
| $b$ | Biting rate | day$^{-1}$ | Brière | $c = 2.02 \cdot 10^{-4}$<br>$T_{min} = 13.35$<br>$T_{max} = 40.08$ | Mordecai *et al.* (2017) |
| $p_S$ | Probability of mosquito infectiousness | | Brière | $c = 8.49 \cdot 10^{-4}$<br>$T_{min} = 17.05$<br>$T_{max} = 35.83$ | Mordecai *et al.* (2017) |
| $\delta_h$ | Intrinsic incubation period | day$^{-1}$ | | 1 / 5.9 | Huber *et al.* (2018) |
| $\gamma_h$ | Human recovery from infectiousness | day$^{-1}$ | | 1 / 5 | Huber *et al.* (2018) |
| $r_h$ | Human birth rate | day$^{-1}$ | | 0 (for fitting);<br>1 / (77 × 365) for 50-year simulations. | |
| $\mu_h$ | Human mortality rate | day$^{-1}$ | | 0 (for fitting);<br>1 / (77 × 365) for 50-year simulations . | |
| $g$ | Number of eggs laid per female | | Brière | $c = 8.56 \cdot 10^{-3}$<br>$T_{min} = 14.58$<br>$T_{max} = 34.61$ | Mordecai *et al.* (2017) |
| $s$ | Probability of egg-to-adult survival | | Quadratic | $c = -5.99 \cdot 10^{-3}$<br>$T_{min} = 13.56$<br>$T_{max} = 38.29$ | Mordecai *et al.* (2017) |
| $d$ | Mosquito egg-to-adult development | day$^{-1}$ | Brière | $c = 7.86 \cdot 10^{-5}$<br>$T_{min} = 11.36$ | Mordecai *et al.* (2017) |

| | | | | | |
|---|---|---|---|---|---|
| | rate | | | $T_{max} = 39.17$ | |
| $1/\mu_v$ | Mosquito life-span (inverse of mortality rate) | days | Quadratic | $c = -1.48 \cdot 10^{-1}$<br>$T_{min} = 9.16$<br>$T_{max} = 37.73$ | Mordecai *et al.* (2017) |
| $p_c$ | Probability of mosquito infection per bite on an infectious human | | Brière | $c = 4.91 \cdot 10^{-4}$<br>$T_{min} = 12.22$<br>$T_{max} = 37.46$ | Mordecai *et al.* (2017) |
| $\delta_v$ | Virus extrinsic incubation period | day$^{-1}$ | Brière | $c = 6.65 \cdot 10^{-5}$<br>$T_{min} = 10.68$<br>$T_{max} = 45.90$ | Mordecai *et al.* (2017) |
| $N$ | Host population size | | | | |
| ${N_v}^{max}$ | Maximum mosquito population size | | | ${N_v}^{max} = N \cdot M$ | |
| $M$ | Ratio of mosquitoes to humans | | | $M = 4$ (from fit) | |
| $k$ | Boltzmann-Arrhenius constant | eV/K | | $8.617 \cdot 10^{-5}$ | |
| $T_0$ | Reference temperature at which carrying capacity is maximised | ℃ | | 29 | |
| $E_a$ | "Activation energy" of the carrying capacity | eV/K | | 0.05 | Huber *et al.* (2018) |
| $\varepsilon$ | Infectious mosquito introduction rate (proportion of the human population) | | | $10^{-5}$ | |

**Appendix S2. Temperature-dependent basic reproductive number $R_0(T)$**

We derived a temperature-dependent formulation of $R_0$ applying the next-generation-matrix approach[3] to the SEIR-SEI model:

$$R_0^{\mathrm{NGM}}(T) = \sqrt{M \frac{b^2(T)\, p_s(T)\, p_c(T)}{((1 + (\mu_h/\delta_h))\,(\gamma_h(T) + \mu_h)\,(1 + \mu_v(T)\,/\,\delta_v(T)))\,\mu_v(T)}}.$$

However, this expression needs to be squared to give the number of humans infected from a single infectious human[4,5], meaning that $R_0 = \left(R_0^{\mathrm{NGM}}\right)^2$. As a result, $R_0$ scales linearly with $M$, the ratio of vectors to humans, which is notably difficult to estimate. In our analyses for estimating the susceptibility index, we assume this ratio to be fixed across states and seasons.

**Table S2:** Grid of parameters used to estimate the initial susceptibility index $S_i$.

| ***Model parameter*** | ***Symbol*** | ***Values*** |
|---|---|---|
| Ratio of vectors to humans | $M$ | $\{0.1, 0.25, 0.5, 1, 2, 3, 4, 5\}$ |
| Initial infected proportion | $I_h$ | $\{0.05, 0.01, 0.005, 0.001, 0.0005, 0.0001\}$ |
| Reporting rate | $a$ | $\{0.001, 0.005, 0.01, 0.05, 0.1, 0.2, 0.3, 0.4\}$ |
| Initial susceptibility index | $S_i$ | [0.05, 0.95] |

**Appendix S3. Results using confirmed cases only**

We used total cases (confirmed plus suspected) in our analyses in the main text. To assess robustness to potential misclassification biases, we repeated analyses using confirmed cases only. Although confirmed cases have their own limitations (e.g., reduced testing during epidemics), results remained very similar, as shown below.

Fig. S2 shows the version of Fig. 2 in the main text using confirmed cases only. The patterns remain almost the same: Pearson correlation coefficients tended to be marginally more negative than with total cases (21 of 24 seasons; mean difference -0.10), though the two case definitions are close for 2024 specifically (confirmed cases: r = -0.71, 95% CI: -0.96 to 0.16; compared to total cases: r = -0.72, 95% CI: -0.96 to 0.09); as with total cases, this interval is wide enough to not be statistically significant. Fitting the same weighted, autocorrelation-corrected trend model described in Methods, the declining trend over time remained similar using confirmed cases: -0.037 per year for Pearson (95% CI: -0.061 to -0.013) and -0.030 per year for Spearman (95% CI: -0.048 to -0.011), compared to -0.059 (Pearson) and -0.044 (Spearman) per year using total cases.

The GAMs fitted to reproductive numbers estimated using confirmed cases only showed some small qualitative differences, as shown in Fig. S4. The temperature at which $R_t$ peaked was essentially the same using confirmed cases (28.3°C, 95% CI: 27.7 to 32.9°C) as using total cases (28.4°C, 95% CI: 27.9 to 28.9°C). At 32°C, $R_t$ estimates were a higher percentile using confirmed cases (75th, 95% CI: 69 to 96; compared to 70th, when using total cases), so reproductive numbers remained comparatively higher at the highest temperatures.

The susceptibility indices inferred from total and confirmed cases were very similar (Fig. S7). The maximum-likelihood parameter set was the same, except for a decrease in the reporting rate from 0.2 to 0.05. The qualitative patterns between temperature and $S_{\mathrm{index}}^{\mathrm{init}}$ remained the same (Fig. S10).

The linear regression models with temperature and lagged final susceptibility indices based on confirmed cases were very similar to those based on total cases (see Table S3 and Fig. S12).

**Table S3:** Out-of-sample metrics for linear regressions. Results for three different out-of-sample metrics, comparing linear regression models based on both total and confirmed cases. Also see Fig. 5 in the main text and Fig. S12.

| **Model** | **Cases** | **Pearson *r*** | **Lin's $\rho_c$** | **RMSE** |
|---|---|---|---|---|
| *T* (lag-zero) | | 0.72 | 0.04 | 0.75 |
| $S_f$ (lag-one) | Total | 0.80 | 0.27 | 0.64 |
| | Confirmed | 0.75 | 0.19 | 0.68 |
| *T* (lag-zero) x $S_f$ (lag-one) | Total | 0.82 | 0.80 | 0.37 |
| | Confirmed | 0.77 | 0.70 | 0.44 |

**Appendix S4: Correlations between temperature and incidence at different spatial scales**

We repeated the temperature-incidence correlation analysis (Figs. 2 and S2) at two finer spatial resolutions: Brazil's 439 SUS health regions (averaging ~12 municipalities each) and its 5,503 municipalities. Health-region totals were built by summing municipality-level cases and population within each region-season and population-weighted mean temperature. At municipality level, a unit was retained only if its mean pre-2024 confirmed incidence exceeded a historical floor (10 and 20 confirmed cases per 100,000 per season for health regions and municipalities, respectively), excluding units too sparse to inform a stable estimate (413/439 health regions, 4,009/5,503 municipalities retained). At each resolution we computed the same spatial-autocorrelation-corrected Pearson and Spearman correlations as Figs. 2 and S2 for every season, summarized their trend with a weighted GAM on the Fisher-z scale, used in place of a linear fit because the decline is concentrated in the most recent seasons rather than spread evenly across the study period. We then also compared the correlation's mean level in the first (2001–2006) and last (2019–2024) quarters of the study period (Fig. S3, Table S4). The sign, trend, and first-to-last-quarter change are all significant at every resolution. The correlation coefficients themselves tend to be nonetheless systematically smaller in magnitude at finer resolution, which can be expected independent of any change in the underlying relationship: correlating means over larger aggregates removes sub-unit variation unrelated to the temperature-incidence association, which can inflate the observed coefficient, and finer units carry far fewer cases (a median of ~2,700 per health-region-season versus ~60 per municipality-season), so both incidence and temperature are more noisily estimated, further attenuating the correlation. That direction, trend, and significance all survive a greater than tenfold increase in spatial resolution despite the shrinkage in magnitude, argues against the state-level result being an artifact of aggregation.

**Table S4:** First quarter (2001-2006) vs. last quarter (2019-2024) nationwide temperature-incidence correlation, by spatial resolution, case type, and correlation type.

| Res. | Cases | Type | 2001-2006 mean *r* | 2019-2024 mean *r* | z | p |
|---|---|---|---|---|---|---|
| State | Total | Pearson | 0.64 | -0.35 | -5.61 | <0.001 |
| | | Spearman | 0.43 | -0.42 | -4.93 | <0.001 |
| | Confirmed | Pearson | 0.49 | -0.39 | -4.89 | <0.001 |
| | | Spearman | 0.31 | -0.44 | -4.24 | <0.001 |
| Health region | Total | Pearson | 0.49 | -0.11 | -5.58 | <0.001 |
| | | Spearman | 0.45 | -0.14 | -5.25 | <0.001 |
| | Confirmed | Pearson | 0.31 | -0.18 | -4.53 | <0.001 |
| | | Spearman | 0.28 | -0.21 | -4.27 | <0.001 |
| Municipality | Total | Pearson | 0.34 | -0.08 | -4.38 | <0.001 |
| | | Spearman | 0.34 | -0.08 | -4.37 | <0.001 |
| | Confirmed | Pearson | 0.22 | -0.11 | -3.57 | <0.001 |
| | | Spearman | 0.22 | -0.12 | -3.59 | <0.001 |

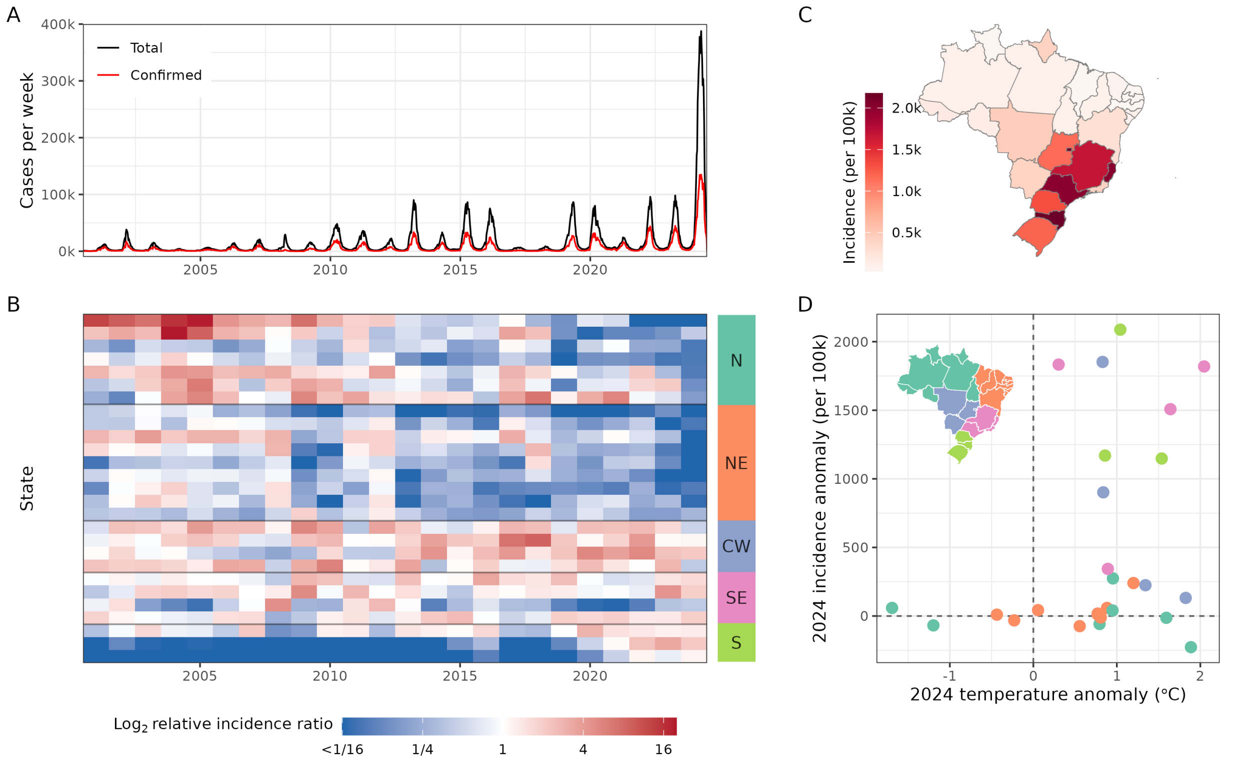


**Fig. S1. Spatiotemporal patterns and drivers of the 2024 dengue outbreak in Brazil.** Same as Fig. 1 in the main text, but using confirmed cases only.

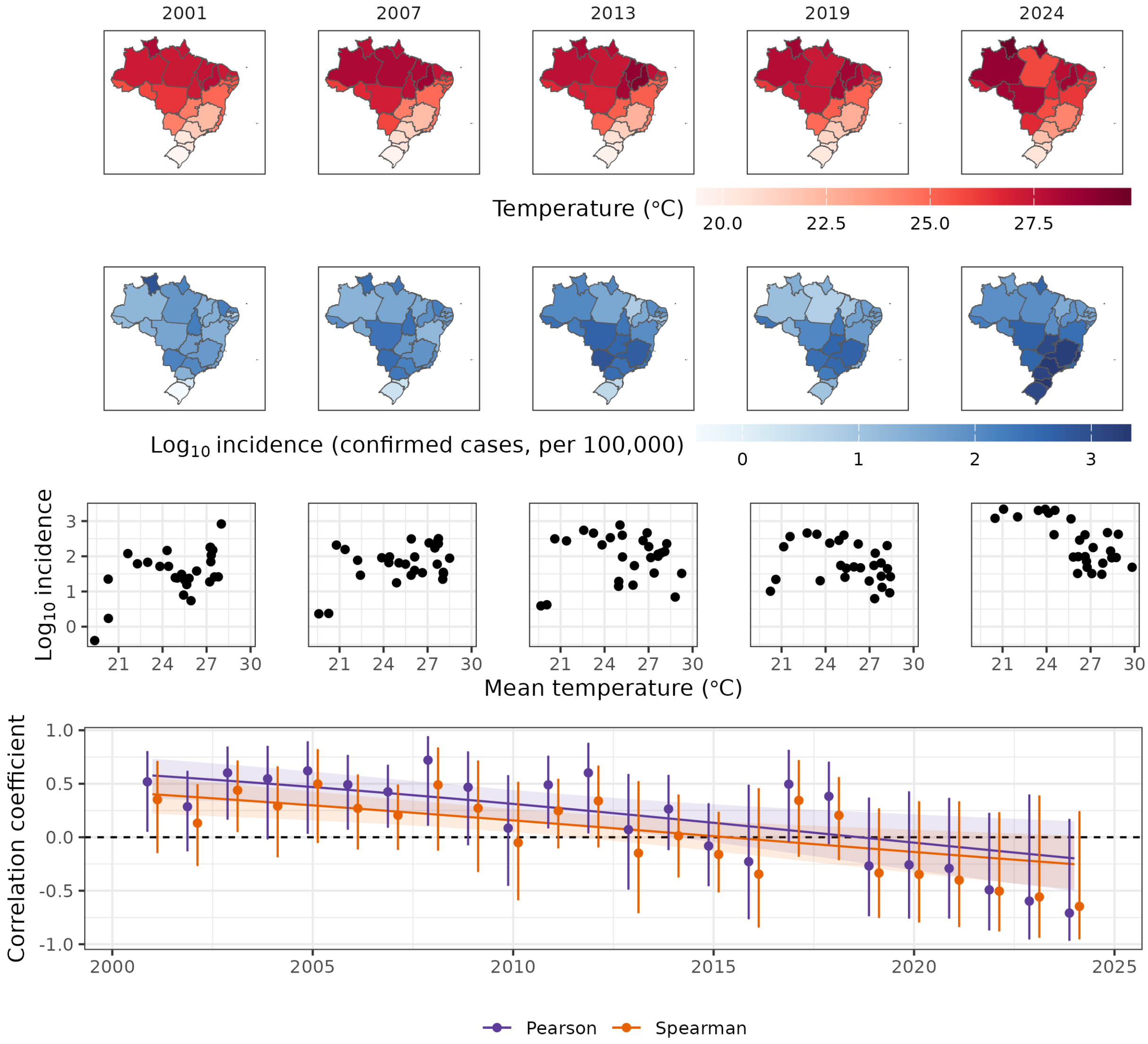


**Fig. S2: Correlations between mean temperature and incidence.** Same as Fig. 2 in the main text, but here using confirmed cases only to calculate incidence.

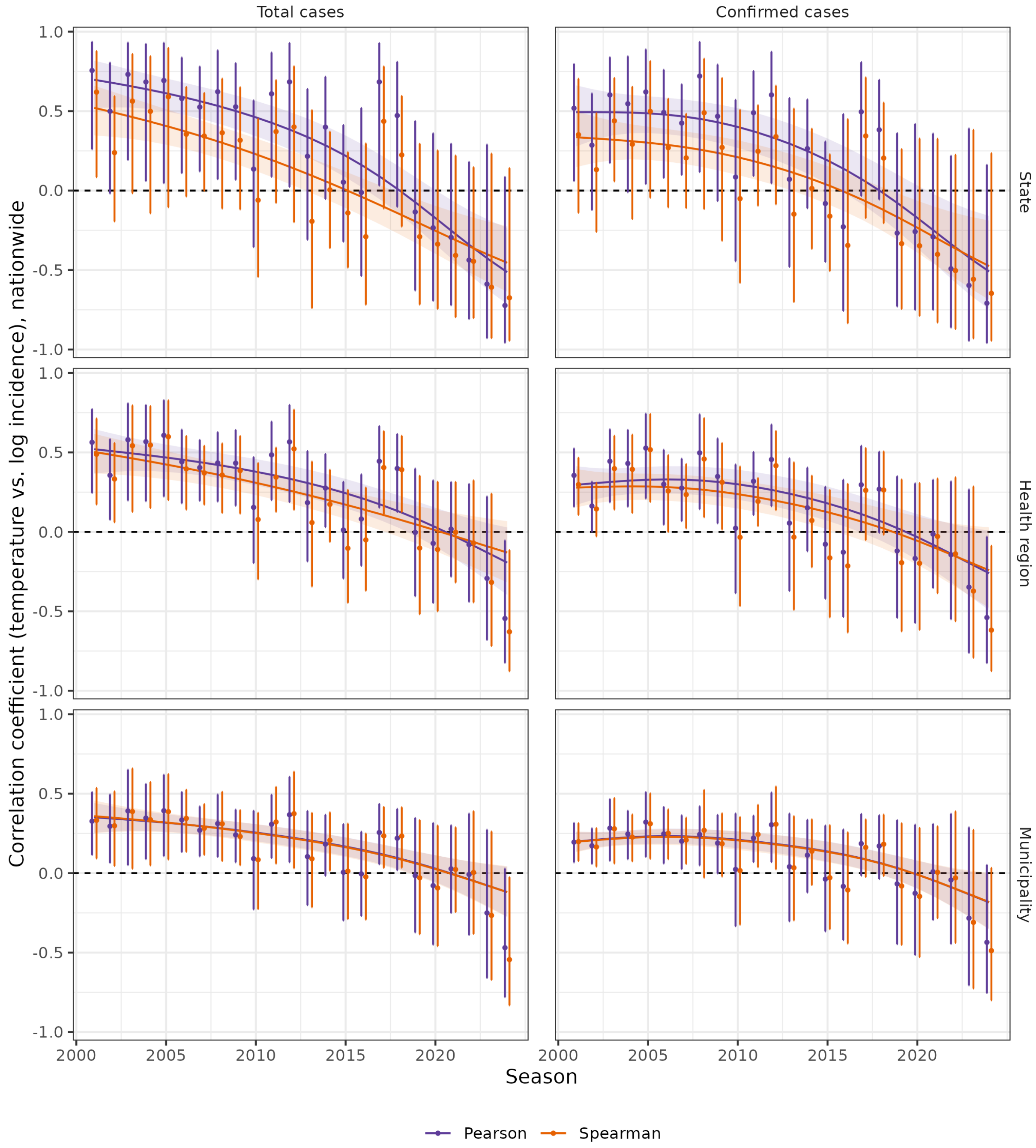


**Fig. S3: Correlation coefficients between temperature and incidence at different spatial scales**. Same as the bottom panel in Fig. 2 in the main text, but using different spatial scales (state level, reproducing the main text panel; health region; and municipality), for both total and confirmed cases.

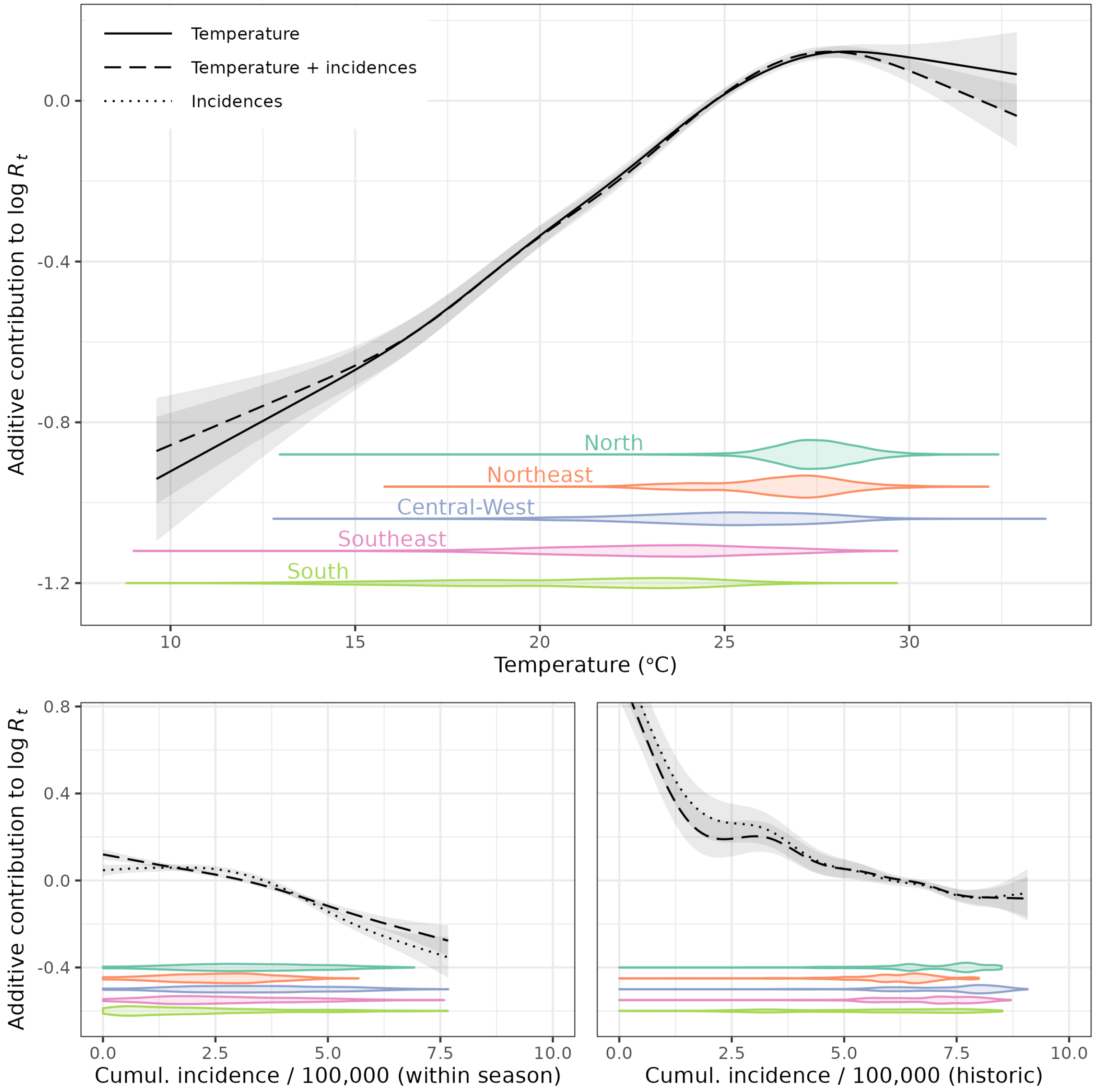


**Fig. S4: Effects of temperature and susceptibility on effective reproductive numbers.** Same as Fig. 3 in the main text, but here using confirmed cases only to estimate effective reproductive numbers, and for the cumulative incidence covariates.

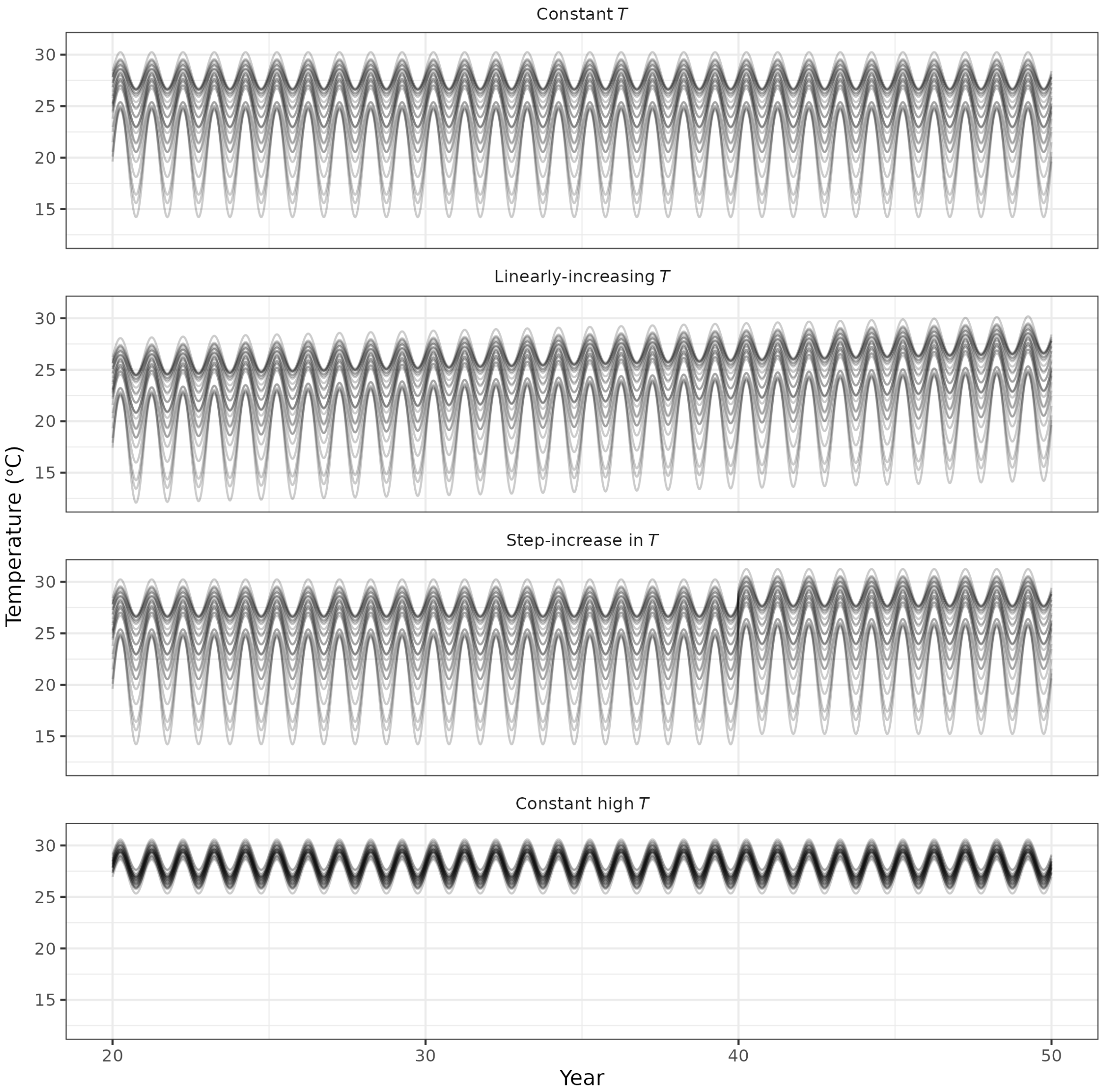


**Fig. S5: Temperature scenarios used in 50-year simulations: constant, linearly increasing, step increase, and constant high temperature**. The four temperature scenarios run for the 50-year simulations. In each scenario, there are 27 lines corresponding to an idealization of each Brazilian state. In the constant temperature scenario, each sine curve has the mean and amplitude that approximately correspond to the observed mean and amplitude in each state (see Fig. S16 below). In the linearly-increasing temperature scenario, the means from the constant scenario are linearly decreased backwards in time. In the step-increase in temperature scenario, the means are increased by one degree in year 40. For the constant high temperature scenario, we assumed all states were similar to the warmer states in Brazil; their mean and amplitude were sampled from narrow normal distributions.

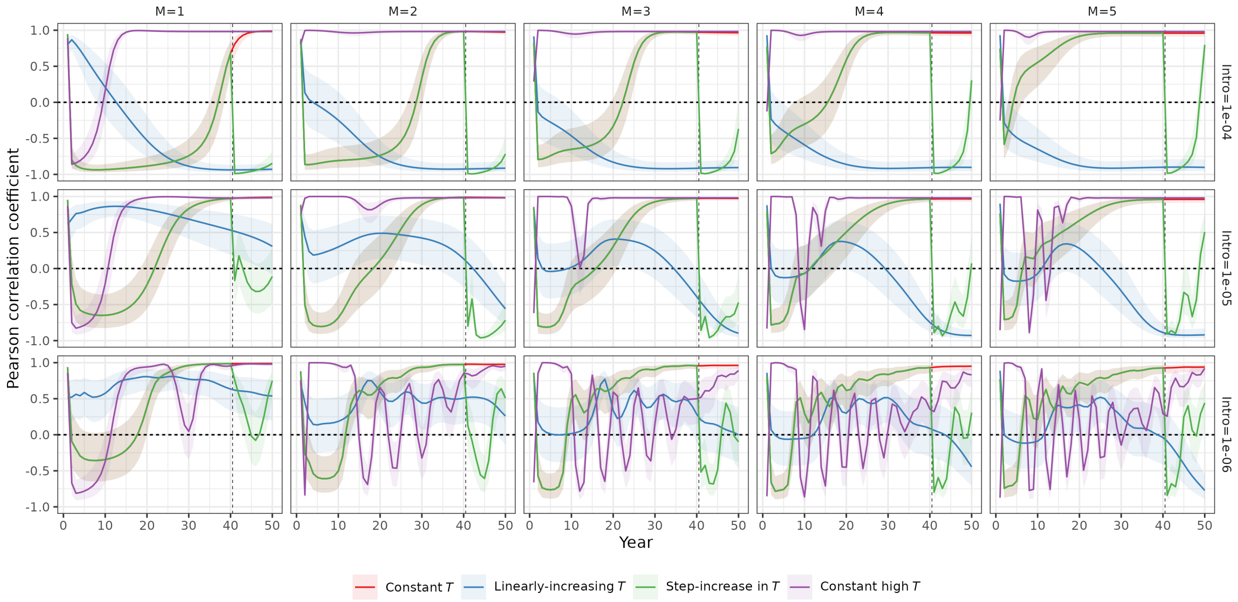


**Fig. S6: Long-term correlations between temperature and incidence across simulation scenarios, mosquito-to-human ratios, and mosquito introduction rates.** The Pearson correlation coefficient between temperature and incidence, for simulations run over 50 years, over the four scenarios described in Methods and Fig. S5, and across five different values of the ratio of mosquitos to humans (columns; *M*), and three different values of constant introduction of mosquitos (rows; see Appendix S1).

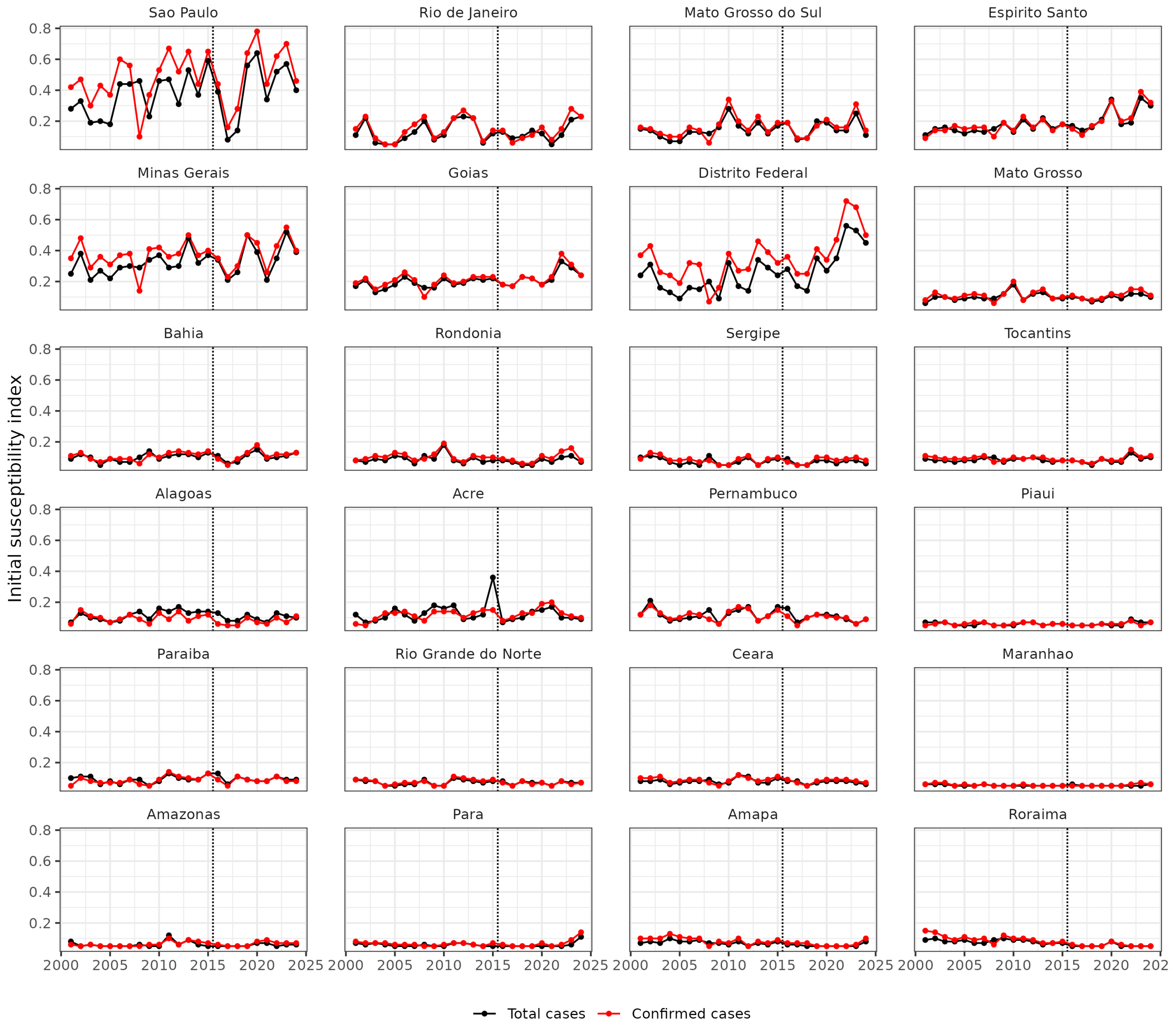


**Fig. S7: Time series of initial susceptibility indices estimated from total case data, 2001–2024.** Time series of the initial susceptibility indices obtained by fitting the SEIR-SEI model to total case data. The maximum likelihood parameters here were: a ratio of mosquitoes to humans $M$ = 4, a reporting ratio $a$ = 0.2 for total cases and $a$ = 0.05 for confirmed cases, and an initial proportion infected $I_h$ = 5x10$^{-4}$. The dotted vertical lines indicate the timing of the Zika outbreak. The black line represents estimates derived from total case data, while the red line represents estimates based solely on confirmed cases.

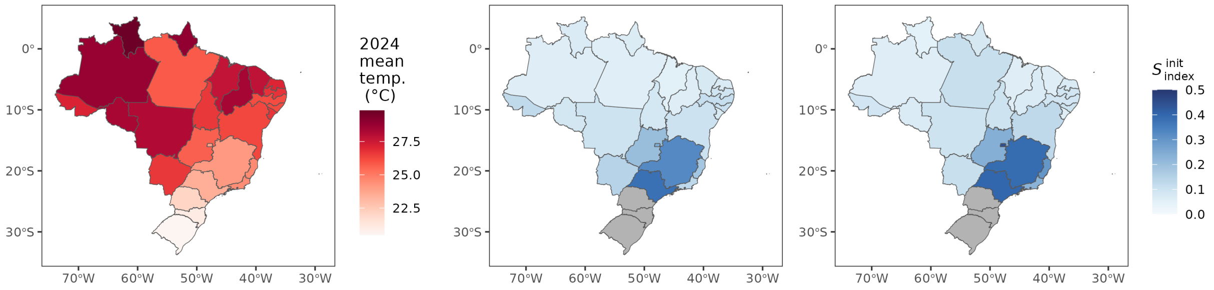


**Fig. S8: Spatial distributions of mean temperature and susceptibility indices.** Spatial distribution of mean temperature for the 2024 season (left) and the initial susceptibility indices estimated using total cases, averaged over the 2001 to 2024 seasons (middle), and for the 2024 season (right; see timeseries in SI Fig. S7).

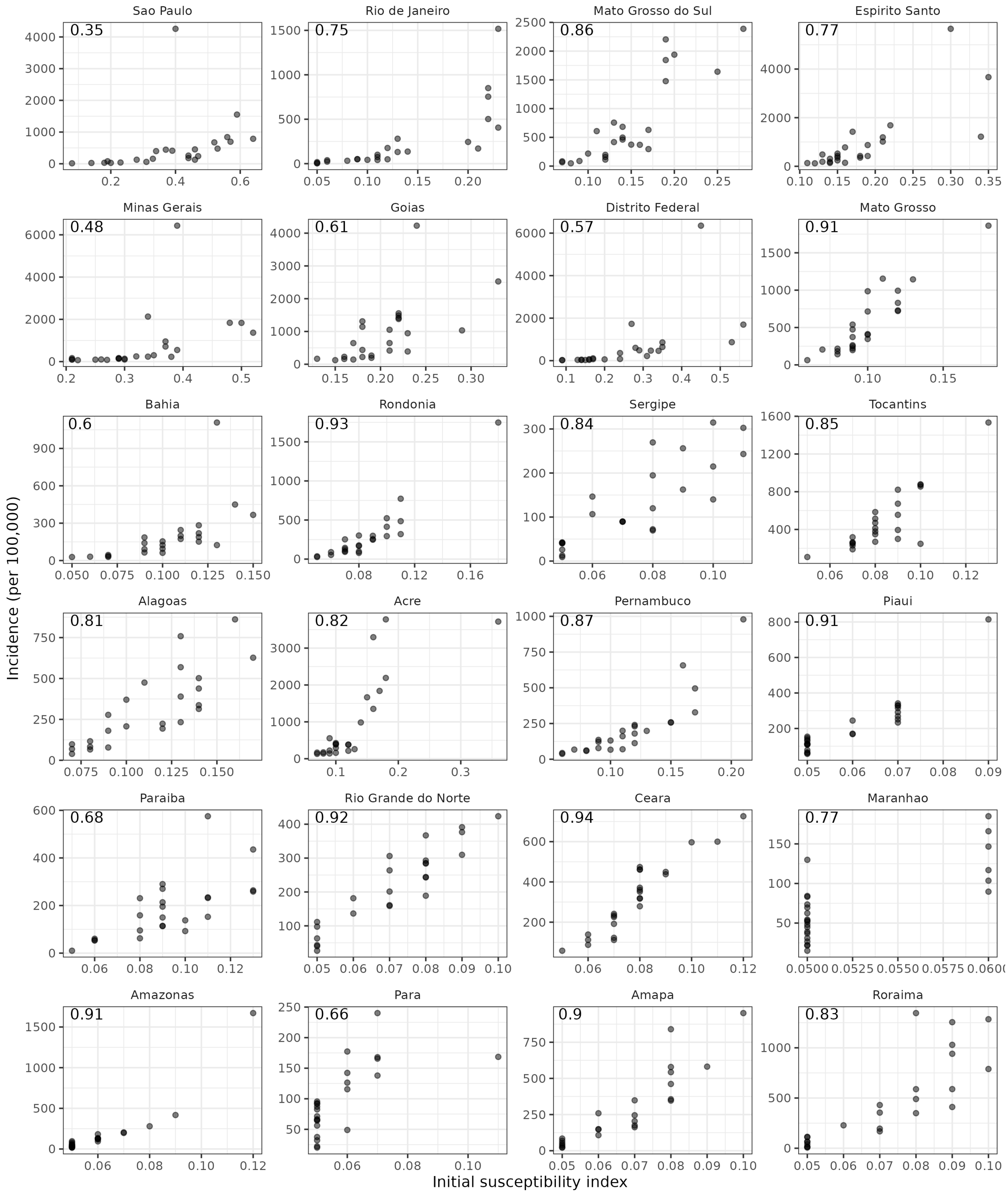


**Fig. S9: Comparison of initial susceptibility indices with incidence.** The initial susceptibility index (based on total cases) as a function of observed incidence (per 100,000, based on total cases). Numbers in the top left corners are the corresponding Pearson correlation coefficients.

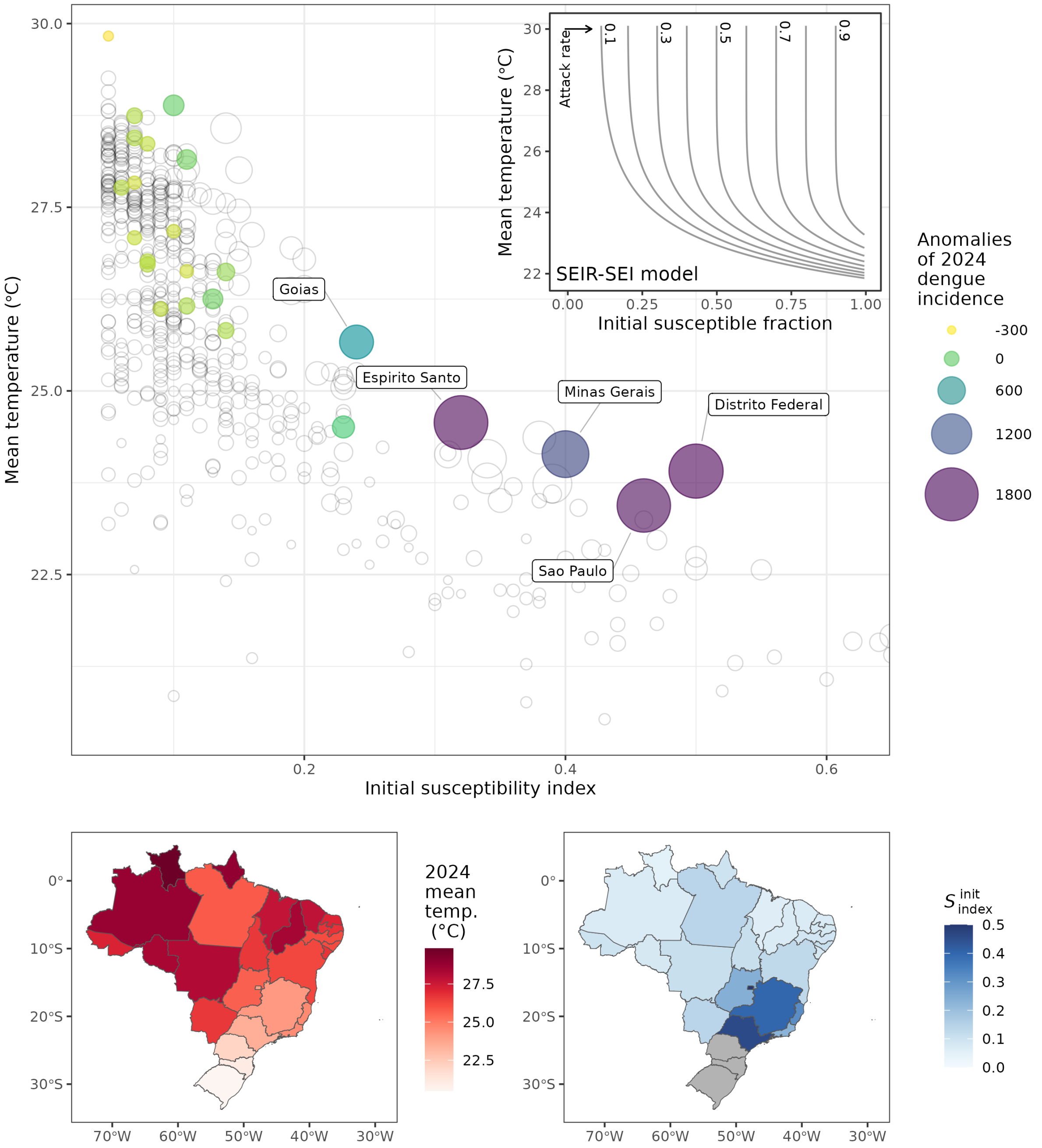


**Fig. S10: Relationship between temperature, susceptibility index, and incidence anomaly using confirmed cases only.** Same as Fig. 4 in the main text, but where we use confirmed cases in the estimation of the initial susceptibility indices.

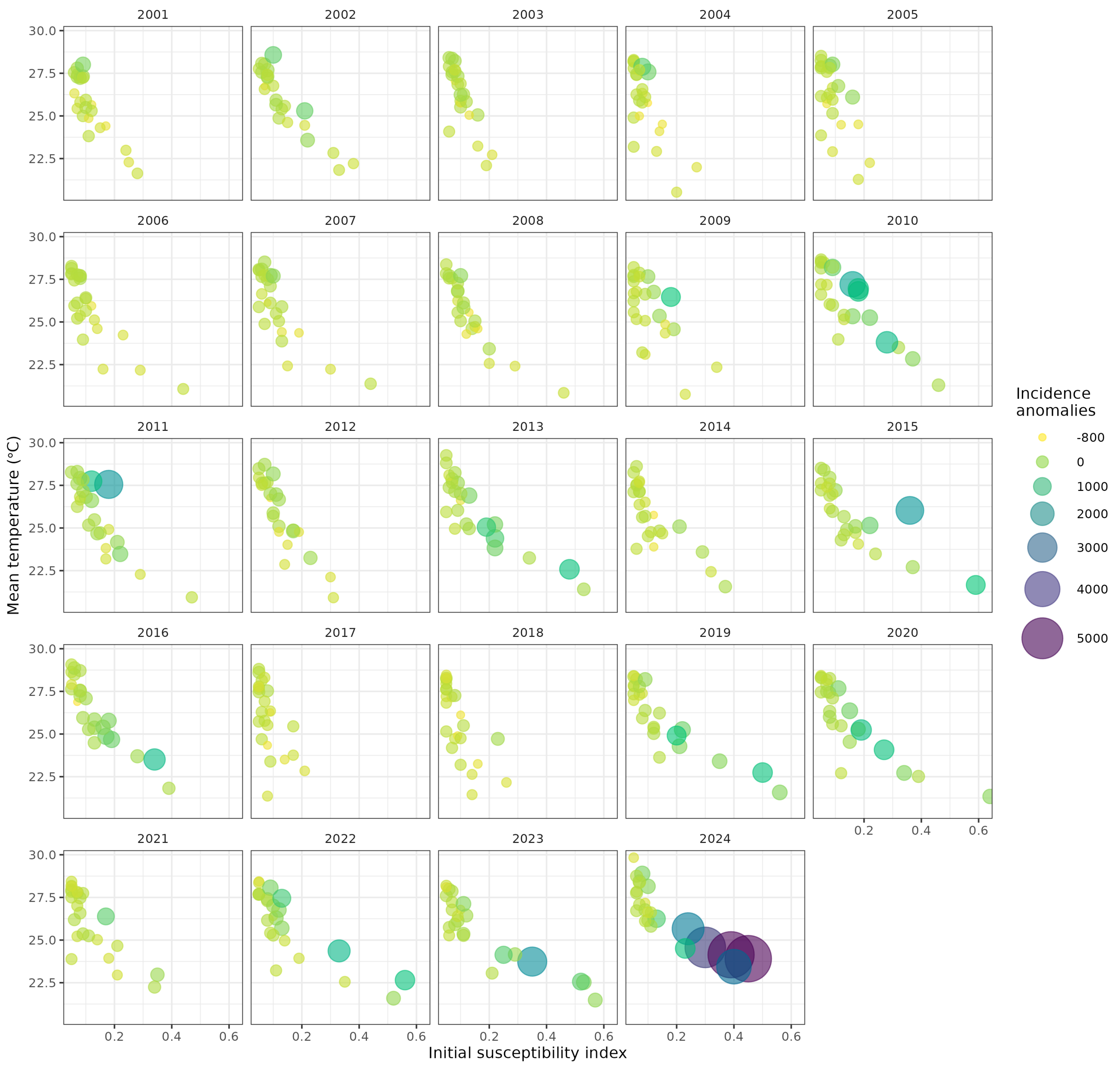


**Fig. S11: The relationship between mean temperatures and initial susceptibility indices.** The incidence anomalies as a function of the initial susceptibility indices estimated using total cases, and mean temperature; as in Fig 4 in the main text, but stratified by season.

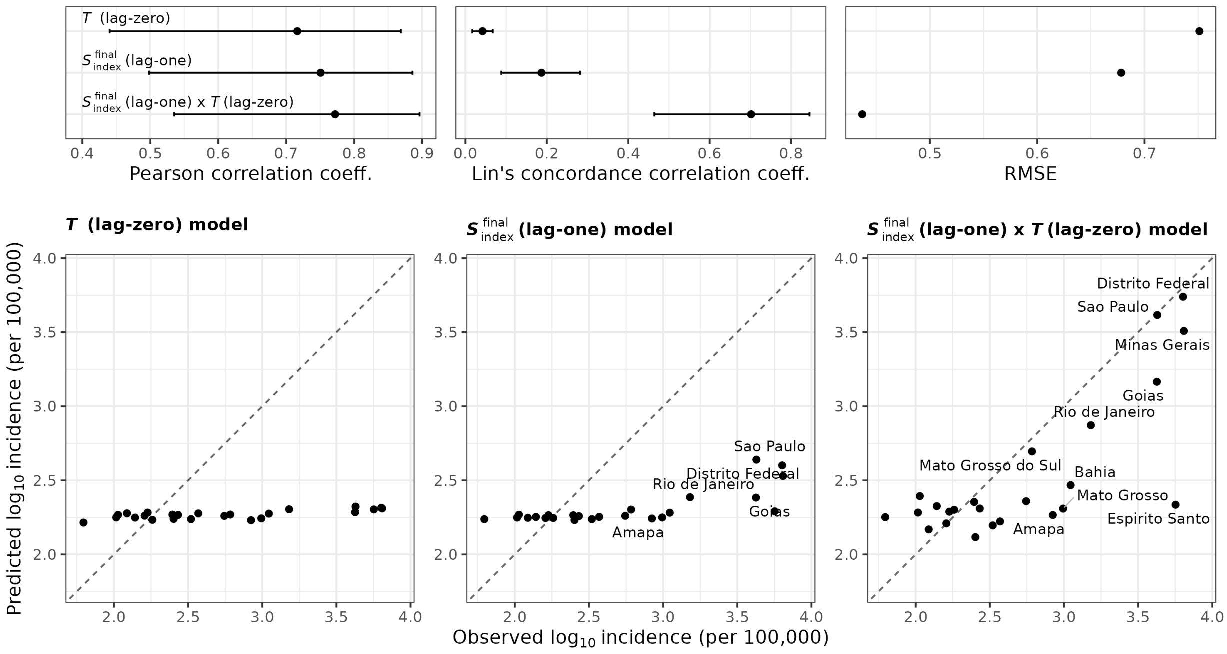


**Fig. S12: Predicting the 2024 outbreak with temperature and susceptibility using confirmed cases only**. Same as Fig. 5 in the main text, but using final susceptibility indices estimated using confirmed cases.

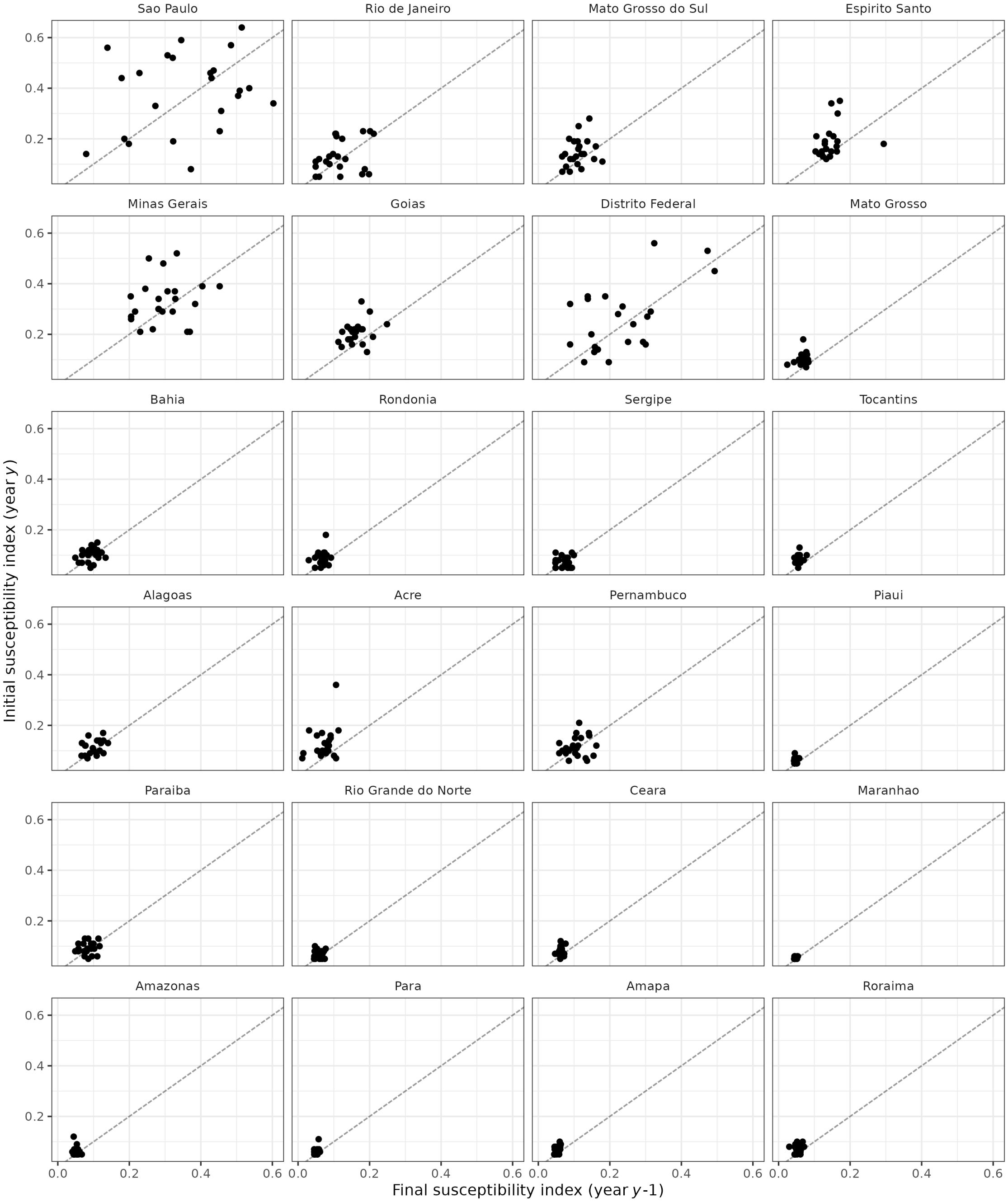


**Fig. S13: Comparison of final and initial susceptibility indices.** Comparison of the final susceptibility for year *y* - 1 (estimated using the SEIR-SEI model, with initial susceptibility set to the initial susceptibility index), with the susceptibility index estimated for the following year *y*. Initial and final susceptibility indices were estimated using total cases. The dotted line is the *y* = *x* line.

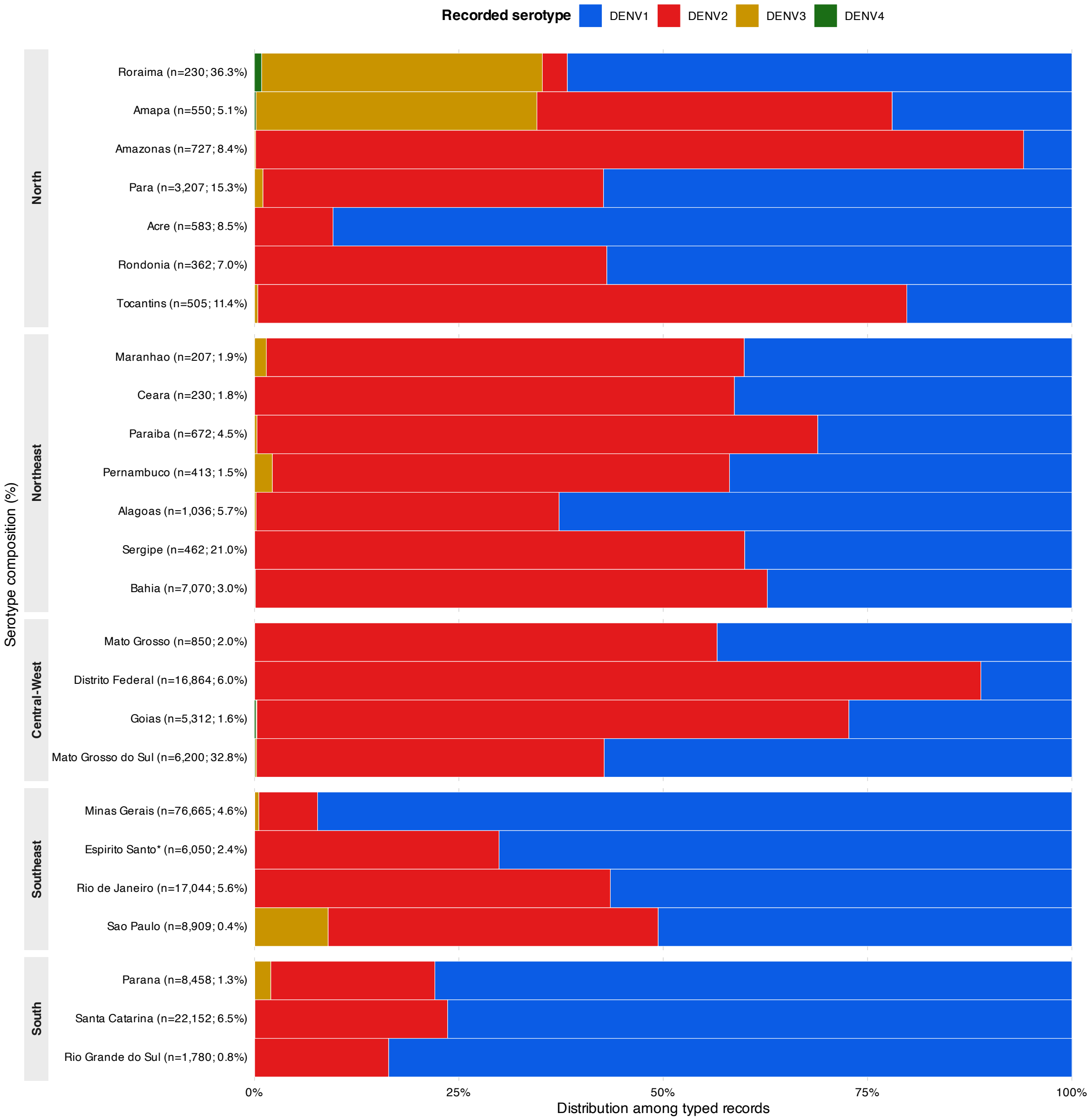


**Fig. S14. Recorded dengue virus serotypes among typed surveillance records in Brazil, 2024.** Stacked bars show the proportional distribution of DENV-1 to DENV-4 among surveillance records with a valid serotype assignment within each state. States with at least 200 typed records are shown and are ordered geographically from North to South. State labels indicate the number of typed records and the percentage of surveillance records with an assigned serotype. Because serotype typing coverage varied among states, the figure should be interpreted as the composition of typed records rather than a direct estimate of statewide serotype prevalence.

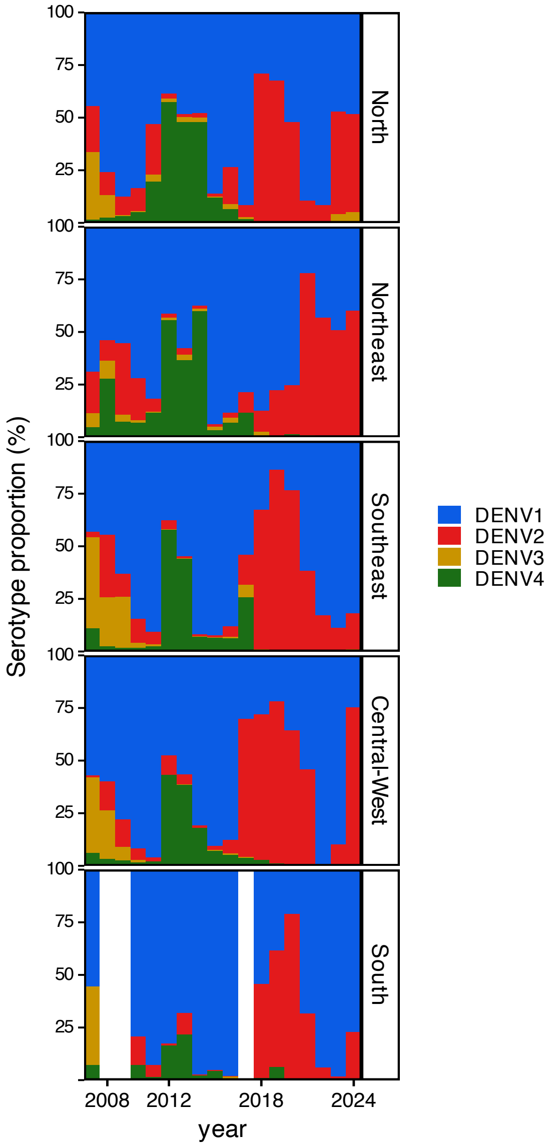


**Fig. S15. Annual dengue virus serotype composition among typed surveillance records by Brazilian region, 2007–2024.** Each stacked bar shows the proportional distribution of recorded DENV-1 to DENV-4 serotypes among surveillance records with a valid serotype assignment within a region and year. Regional-year combinations with fewer than 50 typed records were omitted because of limited sample size. Serotype data were unavailable for 2000-2006. These distributions represent the composition of typed surveillance records and should not be interpreted as population-level serotype prevalence.

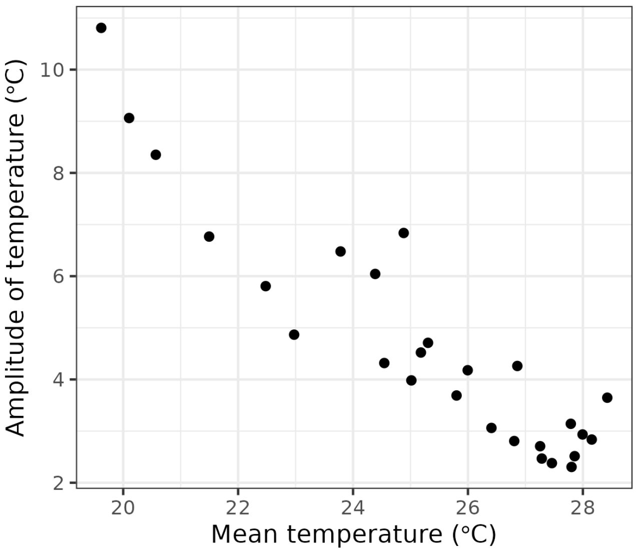


**Fig. S16: Relationship between mean temperature and temperature seasonality (amplitude) across Brazilian states.** The amplitude of temperature (approximated from the standard deviation of temperature) as a function of mean temperature, for each of the 27 states.
